# HIV Transmission Dynamics in Greater Mexico City are Shaped by Dense Spatial Mixing

**DOI:** 10.64898/2026.05.26.26354122

**Authors:** Marina Escalera-Zamudio, Eduardo López Ortiz, Claudia García Morales, Erika Cruz-Bonilla, Shaday Guerrero Flores, Steven Weaver, Margarita Matías Florentino, Daniela Tapia Trejo, Vanessa Dávila Conn, CIENI-CEC Consortium, Maribel Hernandez, Simon Dellicour, Tetyana I Vasylyeva, Joel O Wertheim, Santiago Ávila Ríos

## Abstract

Understanding HIV transmission in densely populated urban settings is essential to mitigate ongoing epidemic spread. We present a comprehensive analysis of recent HIV transmission dynamics in Greater Mexico City, one of the world’s largest metropolitan areas comprising Mexico City and neighbouring municipalities of the State of Mexico. Drawing from over 7,000 complete *pol* gene sequences representing around 50% of new cases reported between 2019 and 2022 within the study region, we reconstructed the transmission network based on pairwise genetic distance. We identified ten large transmission clusters exhibiting sustained growth up to the most recent sampling period. We further analysed paired genetic and high-resolution human mobility data using an integrated phylogeographic approach. We observed a heterogeneous pattern of viral spread across the region, supported by an extensive mixing at a wider geographic scale. Across Greater Mexico City, displaying a high population density, HIV transmission is minimally spatially constrained, a pattern likely fuelled by intense human mobility. Thus, population movement weakens isolation by distance in large urban areas even for a chronic infection that is sexually and vertically transmitted. We demonstrate the value of integrating large-scale genetic, epidemiological, and mobility data to resolve contemporary HIV transmission dynamics in densely populated urban settings.

## INTRODUCTION

HIV transmission dynamics are determined by demographic structure, social behaviour, and sexual networks that shape virus diffusion across spatial and temporal scales^1–3^. HIV spread across urban settings is also closely linked to the timing, frequency, and scale of population-level movement^4^. Understanding the broader determinants underpinning HIV transmission in densely populated urban areas is critical for informing effective public health interventions.

Nearly 400,000 people are estimated to be living with HIV in Mexico^5^, the nation with third greatest burden in the Americas^6^. Over the last four decades, Mexico has experienced a widespread epidemic driven by HIV-1 subtype B^7^. Since 2015, national treatment and prevention programs have successfully reduced prevalence in the general population to a stable 0.3%, with a consistent decline in new cases^5,8,9^. Nonetheless, HIV in Mexico disproportionately affects men who have sex with men (MSM), where prevalence estimates reach up to 11%^10^.

The HIV epidemic in Mexico remains largely urban-centric, with up to 80% of its total population now residing in urban settings^11^. Greater Mexico City encompasses 16 boroughs belonging to Mexico City (named here CDMX) and 59 adjacent municipalities from the State of Mexico (named here MEX)^12^. As one of the world’s biggest metropolitan areas, it encompasses around 8,000 km^2^ and a large and dense population approximating 22 million^11^. Up to 2025, Mexico reported approximately 19,000 new HIV cases annually^5^, with 25% of these registered in Greater Mexico City^13^, underscoring its central role as a major transmission hub.

Recent demographic changes in the country include an increased cross-borders migration^14^ and intensified human mobility, particularly evident in Greater Mexico City^15^. Such transformations have substantially impacted the transmission dynamics of recent and ongoing viral epidemics in the country, exemplified by shifting patterns of SARS-CoV-2 and HIV transmission across international borders^16,17^. In an era of increasing human mobility, Mexico’s urban-centric HIV epidemic provides a valuable opportunity to investigate how recent social transformations—specifically, how far, where, and how frequently people move—contribute to the emergence, establishment and spread of transmission clusters.

The ‘Centre for Research in Infectious Diseases’ at the National Institute of Respiratory Diseases (CIENI-INER, Ministry of Health Mexico)^18^ is a WHO-accredited national reference laboratory leading genomic surveillance of HIV in Mexico since 2016. CIENI-INER works in close collaboration with Clinica Especializada Condesa (CEC, Ministry of Health Mexico)^19^, a public institution that provides free diagnostics and follow-up care for people living with HIV not eligible for the social security system (IMSS and ISSSTE, Ministry of Health Mexico)^20^, being the largest HIV clinic in Latin America. Together, CIENI-CEC conduct routine baseline HIV genotyping to monitor antiviral resistance, centralising up to 70% of newly diagnosed cases reported across the metropolitan area. Official data released by the National Epidemiology Directorate (DGE, Ministry of Health Mexico)^9,21^, which also includes cases reported by IMSS and ISSSTE ^20^, has historically recorded higher absolute case counts and incidence rates in CDMX. However, since 2017, there has been a shift in relative case counts, with higher numbers now reported from MEX^21^.

To characterize the HIV epidemic in Greater Mexico City, we leveraged CIENI-CEC surveillance efforts to access a comprehensive dataset of paired genetic, demographic and epidemiological data. We further used a phylogeographic approach linking genetic and high-resolution human mobility data to assess the drivers of viral spatial dynamics. We find that geographic distance only weakly constrains genetic distance, with viral transmission driven by extensive mixing. Mobility analyses also revealed a heterogeneous spread across geographic regions linked to transmission clusters. By providing insights into the drivers of recent HIV transmission dynamics in one of the world’s largest cities, we present a data-driven analytical framework to investigate HIV spread in large cities with changing demographic structures and social behaviours associated with dense populations.

## RESULTS

### The HIV epidemic in Greater Mexico City

As part of the national drug resistance monitoring effort (**Supplementary Text 1 and Supplementary Fig. 1)**, the CIENI-CEC dataset comprised 9,338 cases sampled nationwide from July 2019 to September 2022. These were filtered to retreive complete HIV *pol* gene sequences (75%; 7,078) from Greater Mexico City only (see Methods section *‘Data access’*). The majority of these (73%; 5,167/7,078) were collected between 2021-2022 from CDMX (70%; 5,106) and MEX (30%; 1,972) (**Table 1, Supplementary Fig. 2**). Sampling mostly represented 30 boroughs/municipalities, markedly skewed towards CDMX, with the Iztapalapa, Cuauhtémoc, and Gustavo A. Madero boroughs reporting the highest case numbers (**Fig.1a**). In contrast to official records comprising surveillance across multiple institutions^9,21^, the CIENI-CEC data displayed higher case counts from CDMX, attributed to a centralised case reporting (**Fig. 1b, Supplementary File 1 - Supplementary Table 1**).

**Fig. 1.**
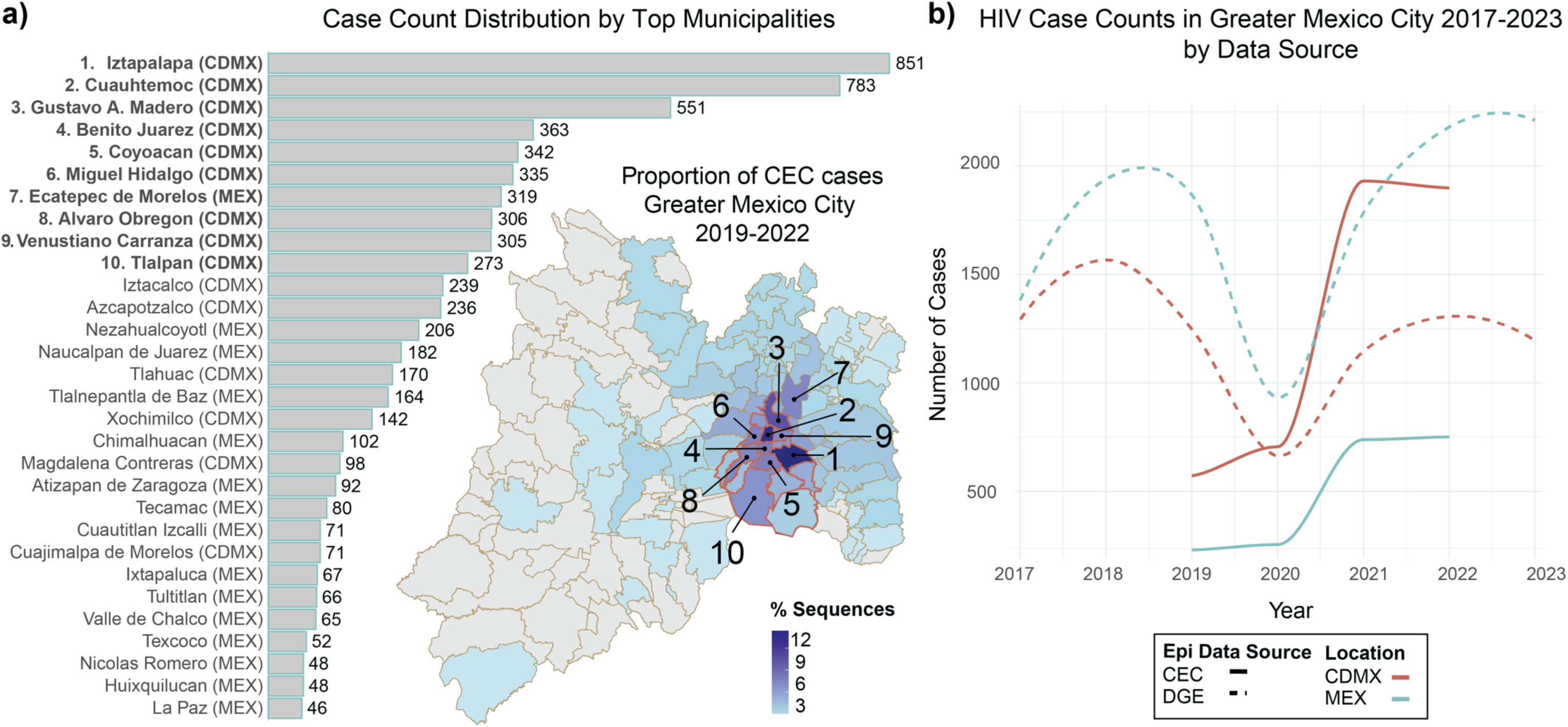
Spatial and temporal distribution of HIV cases in Greater Mexico City. a) Distribution of HIV cases sequenced by CIENI-CEC efforts across Greater Mexico City (Greater Mexico City) between 2019 and 2022. Bars indicate the total number of cases per borough/municipality, ranked by case count. The inset map shows the proportion of CIENI-CEC sequences per borough/municipality, with colour intensity indicating percentage contributions. Numbers on the map correspond to the top ten borough/municipalities with the highest case counts. b) Annual HIV case counts in Greater Mexico City from 2017 to 2023, stratified by data source (CIENI-CEC vs. national epidemiological data) and by location (Mexico City: CDMX, and State of Mexico: MEX). Solid lines indicate values for the CIENI-CEC data, while dashed lines represent national epidemiological data.

**Table 1.**
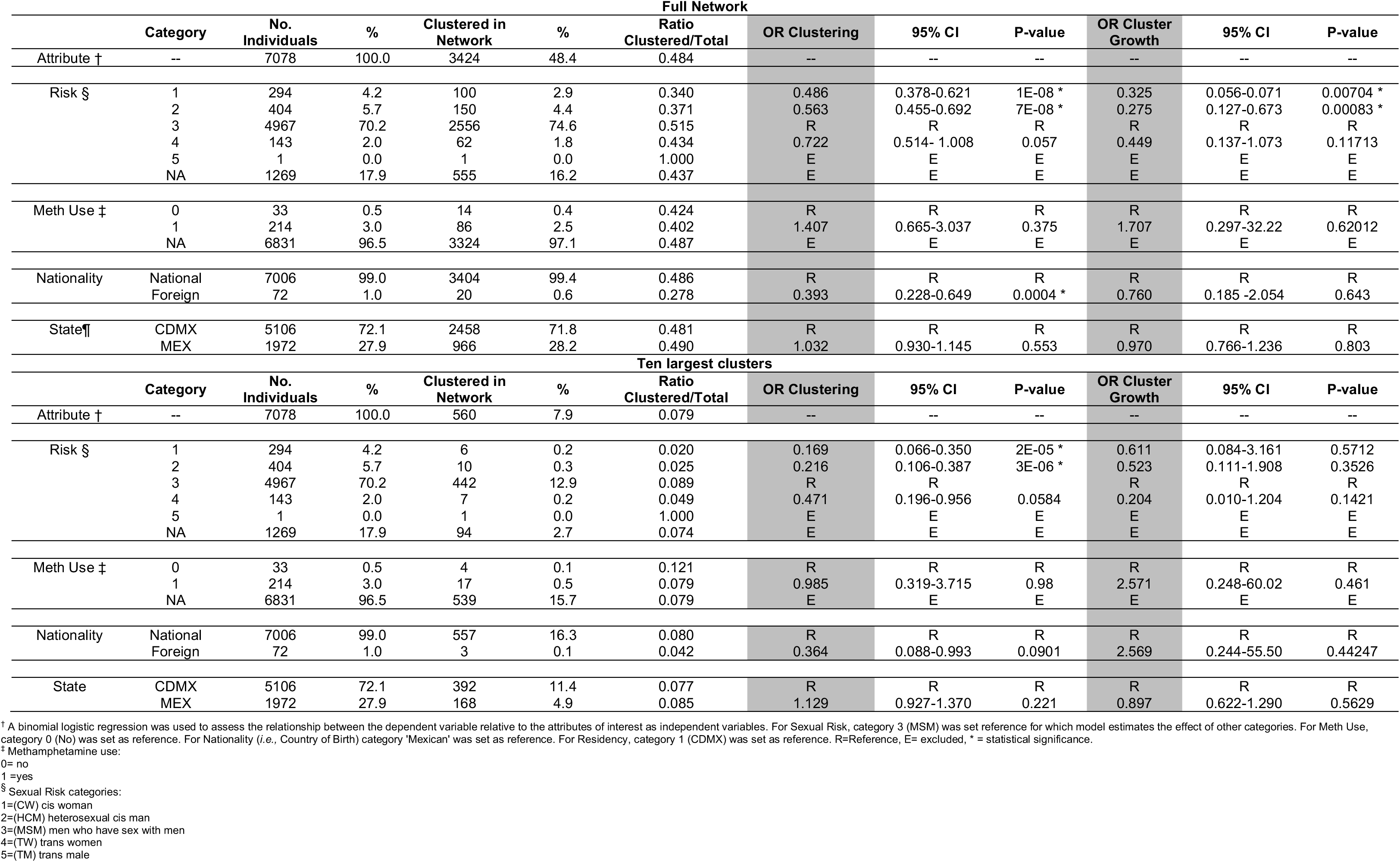
Demographic attributes as predictor of cluster membership and growth.

### An extended HIV transmission network

Drawing from the complete *pol gene* sequence data from Greater Mexico City, we reconstructed the full HIV transmission network based on pairwise genetic distance^22^. Within the network, clusters are defined as a connected component, with nodes representing individual sequences, and edges the genetic distance between sequences under a specified threshold. Applied to CIENI-CEC dataset, we estimated an optimal threshold of 0.015 substitutions per site for the complete *pol* alignment, comparable to that established for the partial 1.4 kb protease-reverse transcriptase region widely used in standard HIV analyses^22^ (**Supplementary Text 1**). Thus, we confirmed that the default configuration for network reconstruction routinely used by CIENI-CEC is correctly calibrated. Extended validation analyses are described in **Supplementary Text 2** and **Supplementary Fig. 3-4**.

The transmission network comprised 990 clusters corresponding to 48% of all sequences (5,873 edges connecting 3,424 nodes. Best-fitting model: Waring distribution, rho = 3.92, 95% confidence interval = [3.76, 4.09], mean cluster size = 3.46 ± 3.02) (**Fig. 2a, Supplementary Fig. 4, Supplementary File 2**). We identified ten large clusters with 15-34 nodes, from which five (clusters number 268, 311, 160, 245 and 11) represented the largest with over 18 nodes (**Supplementary File 1 - Supplementary Table 3**). These ten large clusters were confirmed to be monophyletic and independent in the phylogenetic trees comprising the full CIENI-CEC data (*n* = 7,078) and extended sampling from the Americas (*n* = 37,365) (See Methods section *‘Phylogenetic inference’*) (**Fig. 2b, Supplementary Fig. 5, Supplementary File 1 - Supplementary Table 4**). Although monophyly is not required to define an epidemiologically relevant transmission cluster, this distinction is important because subsequent phylogeographic analyses are conducted on phylogenetically defined clades (that may encompass multiple transmission clusters).

**Fig. 2.**
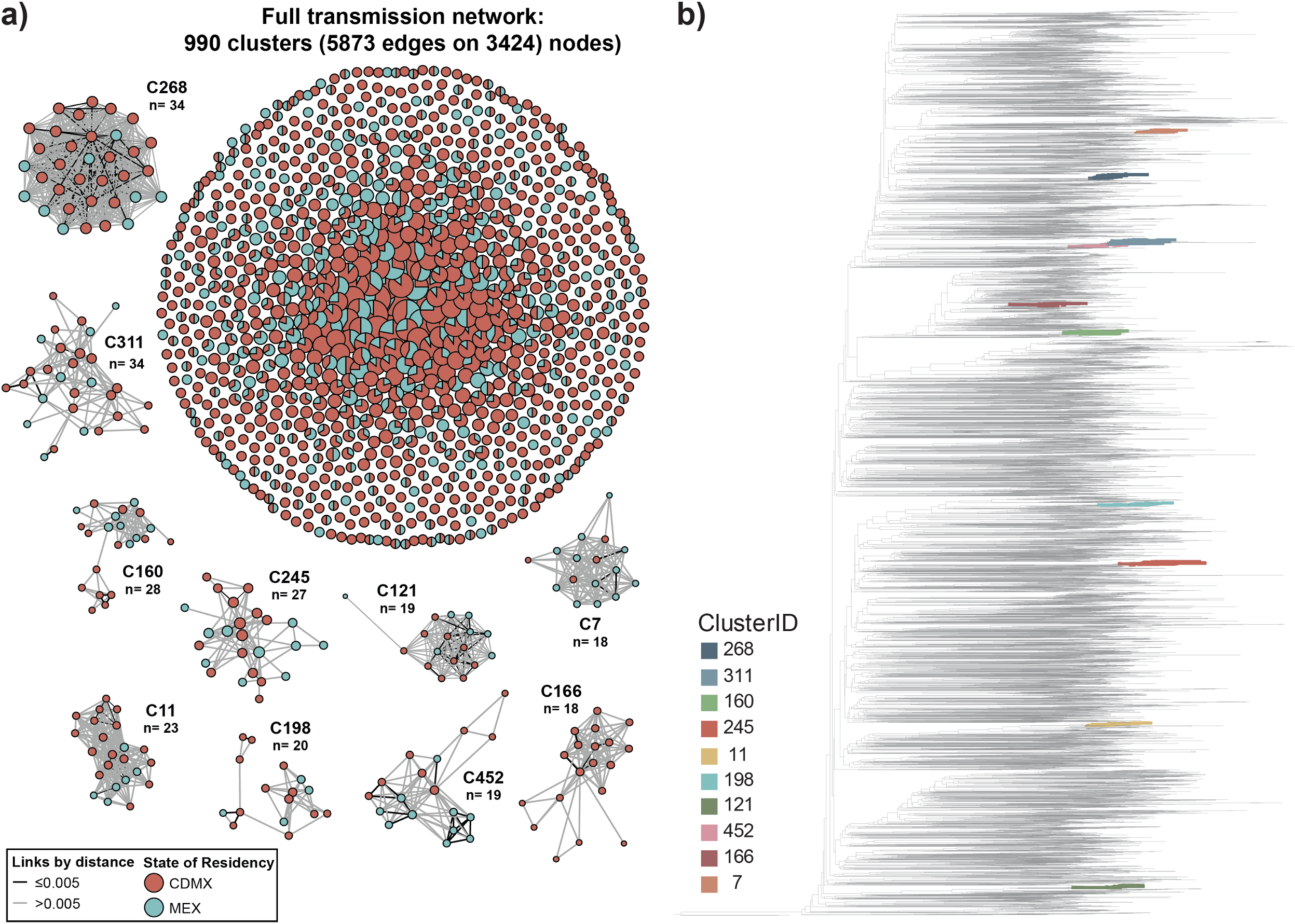
The HIV transmission network for Greater Mexico City. a) The HIV transmission network for Greater Mexico City was reconstructed using HIV-TRACE under a genetic distance threshold of 0.015 substitutions per site. The full network comprises 990 transmission clusters (5,873 edges across 3,424 nodes). Nodes represent individual sequences coloured by state of residence (CDMX in red; MEX in teal). Edges represent putative transmission links inferred from pairwise genetic distances, with line shading indicating genetic divergence (lighter grey representing higher genetic distances, whilst darker lines representing lower genetic distances). Insets show the ten largest transmission clusters (n ≥ 18), illustrating internal connectivity and spatial structure according to sampling locations. b) Maximum-likelihood phylogenetic tree inferred from 7,078 CIENI-CEC sequences from Greater Mexico City. Branches corresponding to the ten largest HIV-TRACE transmission clusters are coloured on the tree, showing that each network-defined transmission chain corresponds to monophyletic phylogenetic clusters.

### Changing demographic structure and social behaviours

Consistent with official epidemiological reports and previous studies^9,13,21,23^, the distribution of demographic variables from the CIENI-CEC data from Greater Mexico City revealed that HIV disproportionately affects young adult males (92%, 6,512/7,078; mean age 31.2 ± 8.9 years) (**Table 1**, **Supplementary Fig. 2, Supplementary File 1 - Supplementary Table 1 and 2**). Men who have sex with men (MSM) represented up to 70% of all cases (4,967/7,078). Self-reported drug use during sexual encounters was frequent (43%, 3,044/7,078), with methamphetamine use increasing over time (0.37% in 2019 to 2.5% in 2022). Although most individuals were nationals (99%, 7,006/7,078), a small proportion of foreign-born individuals was observed (72/7,078), with increasing numbers recorded between 2020 (5 sequences) and 2021 (13 sequences). Most foreign-born individuals came from Colombia (8 sequences) and Venezuela (7 sequences), with other countries in the Americas representing less than 0.1%.

### Asymmetric connectivity within transmission clusters

We then evaluated assortativity within the network, defined as the tendency of nodes to connect to those sharing similar attributes^24^. Assortativity was quantified using the degree-weighted homophily (DWH) ^25^, with values close to 0 indicating random mixing, values close to 1 indicating assortativity (perfect *like-with-like* clustering), and values near −1 indicating disassortativity^24^. DWH indices were computed for four attributes relevant to this study representing a changing demographic structure and social behaviours, and with data availability >1%: methamphetamine use (214 sequences, 3%), nationality (foreign-born individuals: 72 sequences, 1%), sexual risk (excluding trans male group with only 1 sequence) and sampling location (CDMX vs MEX) (see **Table 1**).

For sampling location, we found DWH values close to zero (−0.007), indicating no assortativity across the network. However, when considering only the ten large clusters, spatial heterogeneity is observed through an asymmetric connectivity between nodes. Nodes from CDMX displayed 80% of all connections within CDMX and 20% with MEX. Nodes from MEX displayed 56% of all connections within MEX and 43% with CDMX (**Fig. 2a, Supplementary Fig. 6**). Thus, frequent links across sampling locations likely mask geographic structure across the full transmission network. For all other variables, DWH values close to zero were observed, with an overlap in the corresponding panmictic ranges largely reflecting random mixing (**Supplementary File 1 - Supplementary Table 5**). For extended results, see **Supplementary Text 3**.

### Ongoing growth of transmission clusters

We then assessed cluster growth over time by applying the Sr>2 metric^26^, a standard heuristic used in HIV molecular surveillance (see Methods section *‘Cluster growth and logistic regression’*). Values above 2 flag rapidly expanding clusters based on their relative growth since their earliest sample date. A total of 6.2% (62/990) of all clusters showed evidence of sustained growth. Of the ten large clusters identified, nine displayed values above 2, and five of above 2.5 (C268 = 4.37, C311 = 3.46, C160 = 2.68, C245 = 3.33, and C11 = 3.35; **Supplementary File 1 - Supplementary Table 6**). Our findings suggest that most large clusters have experienced sustained growth over time and across sampling locations, in agreement with the spatial heterogeneity observed.

We further assessed whether the key attributes (*i.e.,* methamphetamine use, nationality, sexual risk group, and sampling location) were associated with cluster growth and structure. Using logistic regression, only sexual risk group was identified as a predictor of cluster growth and structure. Relative to MSM, other sexual risk categories showed lower odds of clustering (44-51% reduction) and of belonging to growing clusters (68-73% reduction, Sr>2; **Table 1**, see **Supplementary Text 3** for extended results). Although MSM comprise the majority of cases, the associations observed derive from odds ratios, and therefore reflect individual-level differences. However, unequal group sizes should be considered when interpreting the magnitude of effects.

### Temporal variations in epidemic growth

To further characterise the temporal growth dynamics of the HIV epidemic in Greater Mexico City, we estimated the joint effective reproductive number (R_e_, the average number of secondary infections generated by an individual at a given time) and the rate of becoming uninfectious (*i.e.,* through treatment, removal from the population, or death). Analyses were conducted across three time periods applied to the ten large clusters under a hierarchical framework (see Methods section ‘*Estimating epidemic growth’*): pre-2017, January 2017 to February 2020, and from March 2020 to December 2022 (corresponding to the timing of the COVID-19 epidemic). Prior to 2017, a R_e_ value of 1.7 (95% HPD: 0.5-3.7) was estimated, followed by a marked increase between 2017 and early 2020 (R_e_ = 3.7, 95% HPD: 1.6-6.7). Consistent with the estimate prior to 2017, after March 2020, R_e_ declined again to remain above the epidemic threshold (R_e_ = 1.7, 95% HPD: 1.2-2.6) (**Supplementary File 1 - Supplementary Table 7**).

Despite temporal fluctuations, sustained endemic transmission of HIV was observed. The rate of becoming uninfectious remained relatively stable across all time periods (0.7-0.9, 95% HPD: 0.1-1.8), corresponding to an average infectious period of approximately 1.1-1.4 years (95% HPD: 0.56-10 years) (**Supplementary File 1 - Supplementary Table 7**). These findings suggest that, during the sampling period, changes in epidemic growth were primarily driven by variation in transmission intensity, such as population-level behaviour, and/or mobility or social disruptions, likely related to the COVID-19 epidemic rather than changes in the rate of diagnosis and treatment^16^.

### Multiple virus introduction events into the study area

To investigate introduction of HIV into Greater Mexico City, we performed a discrete phylogeographic analysis (see Methods section ‘*Discrete phylogeographic analysis’*). We identified a total of 1,958 introduction events into Greater Mexico City, including 497 clades comprising ≥3 samples (**Supplementary Fig. 7, Supplementary File 1 - Supplementary Table 4**). Our results support that the HIV epidemic in Mexico is continuously shaped by multiple, asynchronous introduction events, followed in some cases by local, sustained transmission^27^. Seeding locations cannot be reliably inferred due to overrepresentation^28^, denoting an important sampling bias towards sequences from Brazil and the US. However, the main aim of this analysis was to identify the phylogenetic clades inputted for the subsequent phylogeographic reconstructions described below.

### A heterogeneous spatial mixing

We further investigated the spread dynamics of HIV lineages within the study area using continuous phylogeographic reconstruction^29,30^. Overall, we observed that viral lineages, (*i.e.,* phylogenetic clades) have broadly spread across Greater Mexico City area, following a centralised, time-dependent diffusion pattern radiating from CDMX outwards (**Fig. 3a**). Despite the overrepresentation of sequences from CDMX, the pattern was consistent across three independent replicate control datasets subsampled according to time period-specific incidence (number of cases per 100,000 inhabitants) respective to each sampling location **Supplementary File 1 - Supplementary Table 1**). Consistent with an asymmetric connectivity observed across transmission clusters, most clades exhibited high spatial mixing denoted by frequent viral transitions across sampling locations, with only a few examples of localised virus diffusion patterns (**Fig. 3b-d, Supplementary Fig. 8**). The largest number of lineage dispersal events were estimated ‘within CDMX’ (6856; 95% HPD = 6776, 6913) compared to ‘within MEX’ (1259; 95% HPD = 1189, 1314). We also identified a higher number of dispersal events ‘from CDMX to MEX’ (1023; 95% HPD = 1004, 1054) compared to ‘from MEX to CDMX’ (385; 95% HPD = 353, 420), further confirmed in the control datasets abovementioned (**Fig. 3e**). While the analysis of the full dataset suggests a potentially higher proportion of dispersal events ‘from MEX leaving MEX’ (0.234; 95% HPD = 0.221, 0.247) compared to those ‘from CDMX leaving CDMX’ (0.130; 95% HPD = 0.128, 0.132), this pattern was not recovered across the control datasets (**Fig. 3f**), indicating sensitivity to sampling structure.

**Fig. 3.**
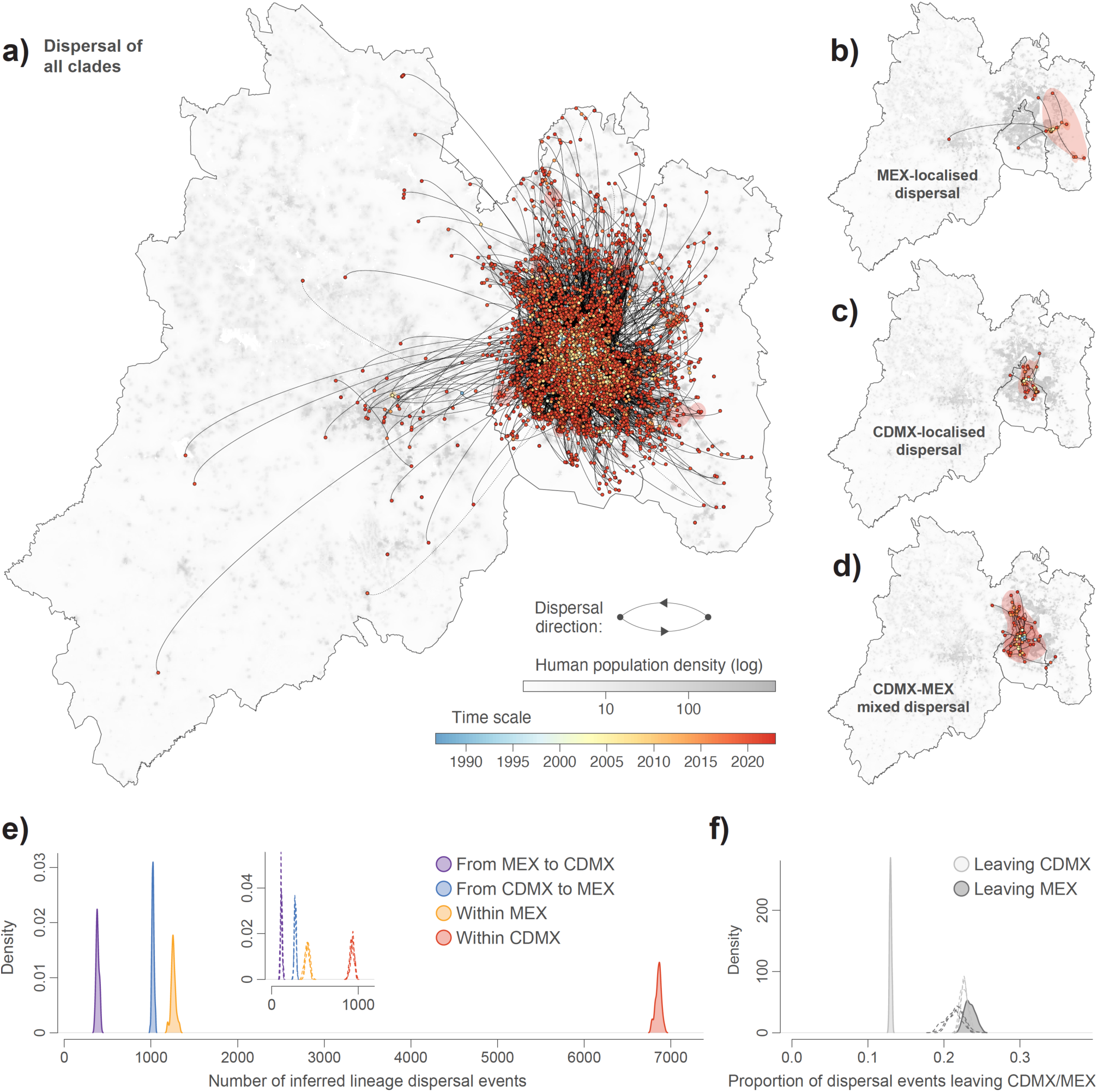
HIV spread across Greater Mexico City. a) Dispersal history of viral lineages as inferred by a continuous phylogeographic analysis conducted along each clade identified as a distinct introduction event into the Greater Mexico City (Greater Mexico City). For each clade, we here map the maximum clade credibility (MCC) tree retrieved from the corresponding continuous phylogeographic inference, with internal and tip nodes coloured according to their estimated time of occurrence and sampling date, respectively. MCC tree branches are displayed as curved arrows indicating the directionality of lineage dispersal events, and shaded polygons also coloured according to time represent 80% highest posterior density (HPD) regions summarising spatial uncertainty of inferred lineage locations for one-year time slices. Background shading corresponds to log-transformed human population density per raster cell. Representative examples of contrasting diffusion patterns observed among individual clades are shown in: b) MEX-localised dispersal, c) CDMX-localised dispersal, and d) mixed CDMX-MEX dispersal. e) Density distributions of the number of inferred lineage dispersal events partitioned by category: within CDMX, within MEX, from CDMX (to MEX), and from MEX (to CDMX). f) Density distributions of the proportion of dispersal events leaving each sampling location (CDMX or MEX) summarised across all phylogeographic clades. In panels e) and f), dashed curves correspond to the results obtained when the analyses are performed on subsampled datasets (see the text for further detail).

### Virus dispersal is weakly spatially constrained

Two viral lineage dispersal statistics — the weighted diffusion coefficient^31^ and isolation-by-distance (IBD) signal^29^ metrics — were estimated for the 497 clades identified as distinct introduction events. The weighted diffusion coefficient measures the average rate of spatial spread of viral lineages through time^31^, whilst IBD corresponds to the relative increase in genetic divergence compared to geographic distance^29^. We observed a limited IBD signal at the metropolitan scale, denoted by a weak correlation between pairwise genetic (patristic) and geographic distances (Spearman’s ρ = 0.269; see **Supplementary Fig. 9** for correlation estimates by clade). Replacing geographic distance by our pairwise mobility metric (see Methods section ‘*Mobility analyses’*) further weakened this association compared to the geographic distance among pairs of sequences whose sampling locations can be connected using mobility data (Spearman’s ρ = −0.141 vs 0.186).

### A heterogeneous spatial spread of transmission clusters

To contextualise the heterogeneous spatial patterns observed, we then analysed human mobility flows within and across Greater Mexico City linked to transmission clusters, based on the residence locations allocated to individual sequences. We observed broadly balanced flows, with a comparable proportion of trips fluctuating over time between January 2020-December 2021 (**Fig. 4a**; for extended results, see **Supplementary Text 3**). The overall temporal distribution of mobility data revealed reduced activity during early 2020, followed by a progressive recovery after early 2021 (**Fig. 4b**), consistent with published reports^15,32^. Mapping mobility flow intensity at a geographic scale revealed substantial movement within CDMX, mostly centralised in the central-western city area, denoted by the Miguel Hidalgo, Cuauhtemoc and Benito Juarez boroughs. High-intensity bidirectional movement was also observed towards neighbouring municipalities in MEX and Toluca City (the capital of MEX) (**Fig. 4c**), in line with well-documented intermunicipal commuting for employment, leisure and services^12^.

**Fig. 4.**
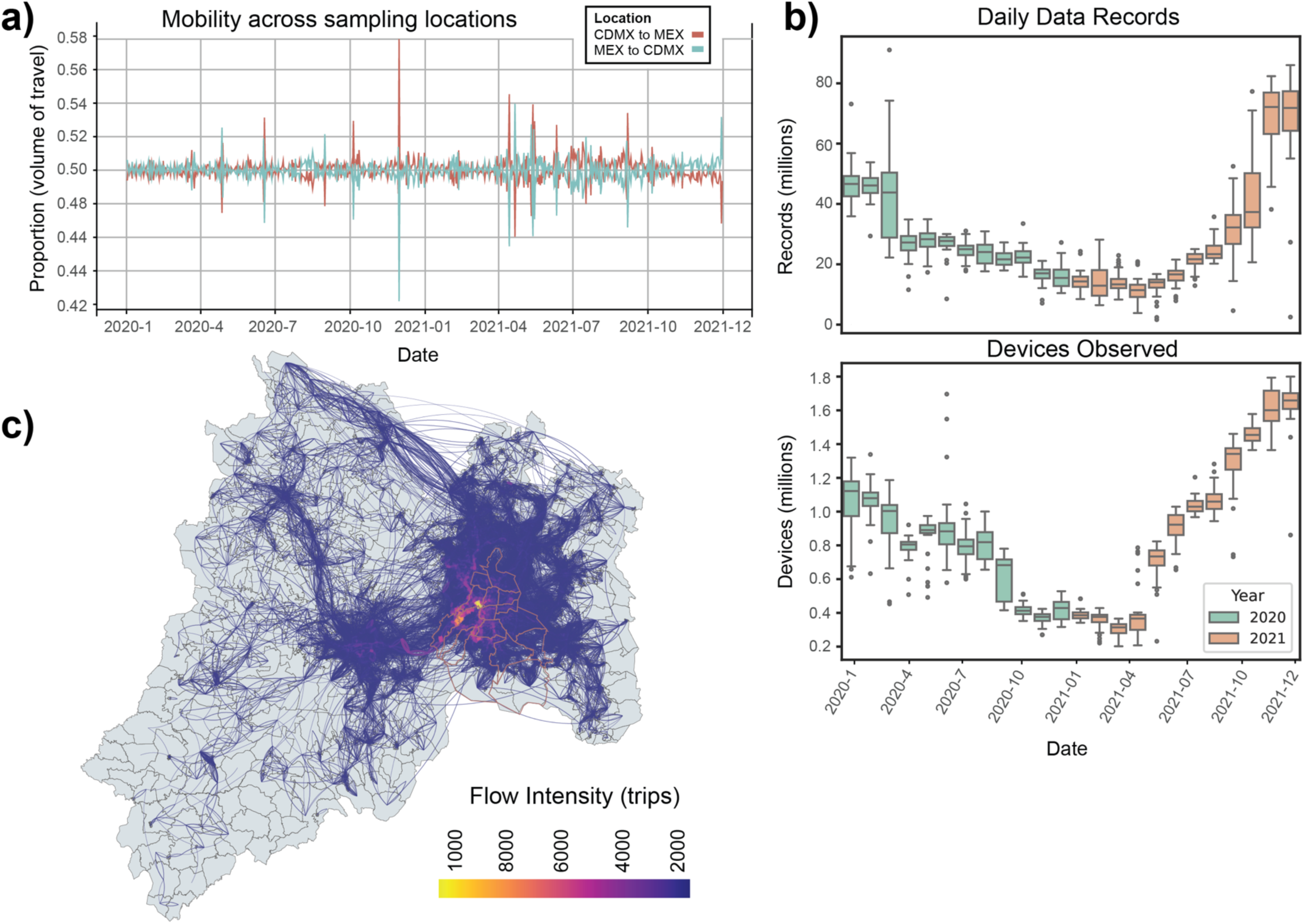
Spatiotemporal structure of human mobility network across Greater Mexico City. a) Temporal dynamics of bidirectional human mobility between CDMX and MEX derived from anonymized mobile device data spanning January 2020-December 2021. Lines represent the normalized proportion of total travel volumes occurring from CDMX to MEX (red) and from MEX to CDMX (teal) per unit of time. Mobility flow fluctuations indicating broadly balanced bidirectional movement across the metropolitan area over time with a marked increase after 2021. b) Temporal distribution of mobility data coverage. Upper panel shows the total number of daily records (millions) captured per month, and lower panel shows the number of active devices observed (millions), stratified by year (2020 in green; 2021 in orange). Boxplots summarise the distribution of daily records aggregated per month, illustrating reduced activity during early 2020 followed by progressive recovery and expansion in 2021. c) Geographical representation of the mobility network across Greater Mexico City reconstructed from aggregated origin-destination flows between AGEBs. Edges represent directed trips between locations, with colour intensity reflecting flow magnitude (number of trips). Only connections with more than two trips are displayed (arc curvature = 0.2). The network reveals dense connectivity within the urban core of CDMX and sustained cross-boundary movement linking CDMX with surrounding municipalities in MEX, consistent with persistent metropolitan-scale population exchange.

Mobility data further support a heterogeneous spatial spread of individual transmission clusters (**Supplementary Fig. 10**). As an illustrative example, C268, the largest and most persistent cluster identified, showed a limited geographic spread, with mobility connections largely confined to CDMX. In contrast, C7, one of the smaller clusters, displayed a wide dispersion range, extending into northern MEX. Finally, we identified mobility hotspots associated with transmission clusters, defined as the top 50 AGEBs (Basic Geo-statistical Area; Area Geoestadistica Basica. See Methods section *‘Mobility analyses*’) receiving the highest cumulative inbound mobility from ‘home’ AGEBs (*i.e.,* residence locations allocated to each sequence). Mobility hotspots were largely concentrated within central CDMX and broadly overlapped with the spatial distribution of the corresponding ‘home’ AGEBs (**Supplementary Fig. 11**). Together, the mobility patterns observed provide a spatial context for the heterogeneous spread of HIV transmission clusters, suggesting that population movement contributes to a weak spatial structuring.

## DISCUSSION

HIV transmission in Greater Mexico City occurs across a highly dense and interconnected urban population, evidenced by a viral genetic diversity only weakly structured by geographic distance. High spatial mixing was observed both within the HIV transmission network and through our phylogeographic reconstructions. Furthermore, mobility data linked to patient residency locations also evidenced a heterogeneous spread of transmission clusters across the metropolitan area. Most studies integrating viral genomics and human mobility have focused on acutely transmitted respiratory viruses, widely explored in SARS-CoV-2^33,34^ and influenza^35^ viruses, demonstrating that transmission dynamics are driven by human movement. However, whether similar drivers shape the disease dynamics of sexually transmitted viruses in large urban settings remained largely unexplored, particularly at this resolution and scale. We demonstrate that human mobility contributes, to some extent, to a heterogeneous spatial spread of HIV across large, densely populated urban areas. Thus, our observations evidence that physical distances, likely overridden by intense population-level movement, do not strongly constrain transmission of this chronic STI.

Quantifying complex drivers of pathogen transmission dynamics remains a long-standing challenge, partly due to the difficulty of integrating multidisciplinary data into analytical frameworks^36^. However, as surveillance systems improve and richer datasets become increasingly available, new opportunities emerge. Combining geospatially resolved genetic data provides a powerful approach for investigating viral spread across spatio-temporal scales^28^. Complementary approaches using human mobility records further enhance this perspective, with clear relevance for public health decision-making^33–35^. In this light, the mobility dataset used here previously enabled the quantification of time-resolved population-level movement trends across Mexico^32^, as well as investigating how non-pharmaceutical interventions reshaped mobility patterns during the COVID-19 epidemic^15,37^.

However, unlike our previous analyses of SARS-CoV-2 genomic data from the region, for which mobility records could not be directly linked to viral sequences^16^, the analyses presented here offer a scalable framework integrating high-resolution mobility and paired viral genetic data into phylogeographic reconstruction. Such an approach could be transferable to other urban hubs with comparable datasets. Nonetheless, given the nature of the mobility metric we used in our study, which mainly captures aggregated population-level movement intensity across locations, a key observation is the reduced magnitude of the association between mobility and viral genetic divergence compared to geographic distance. This distinction suggests that the current metric captures less of the underlying genetic structure.

In the context of our study, the timing of the COVID-19 epidemic is particularly relevant for interpreting local HIV transmission dynamics in Greater Mexico City. Our sampling period encompasses the national lockdown implemented during 2020^16^, after which the mobility network across the metropolitan area was structurally reshaped^37^. Despite fluctuations in human mobility, HIV transmission remained resilient, with R_e_ estimates persisting above the epidemic threshold throughout the sampling period. Thus, our findings suggest that short-lived disruptions in human movement alone are insufficient to disrupt HIV transmission. Although our analyses mainly suggest that changes in epidemic growth were primarily driven by variation in transmission intensity, disruptions to HIV-related healthcare services during the COVID-19 epidemic may still have contributed to local transmission. However, such effects remain to be assessed, as these variables were not explicitly modelled.

Thus, future analyses integrating demographic and behavioural data could help assess how large social events, changing population structure, and varying contact patterns contribute to temporal fluctuations in HIV transmission dynamics. For example, comparing patient residence with locations of sexual encounters and app use may help disentangle where transmission occurs and enable targeted interventions. However, this requires careful consideration of privacy, traceability, and potential stigma associated with identifiable locations. Moreover, resolving the impact of short-lived events remains limited by the evolutionary timescale of HIV over short epidemiological windows.

The spatial and demographic patterns observed are likely linked to recent social transformations occurring across the study region. For example, trends reflecting evolving behavioural and social dynamics, such as increased migration and methamphetamine use, may have important implications for HIV transmission and should remain under close monitoring^38–40^. In parallel, spatial trends may also be linked to recent structural drivers of demographic redistribution. As an example, across Greater Mexico City, rising housing costs in the city have increasingly displaced residents to peripheral municipalities, while employment, healthcare, and social activities remain strongly centralised^41^.

Within the reorganised urban landscape of Greater Mexico City, our analyses suggest a potentially higher proportion of dispersal events ‘leaving MEX’ and ‘within MEX’ are relevant. This potential shift, together with the identification of mobility hotspots linked to transmission, could highlight areas of public health relevance where interventions should be prioritised. However, the extent to which this reflects a change in viral spread dynamics or redistribution of existing infections remains to be resolved as more genetic data becomes available. In the case of dispersal events ‘leaving MEX’, further accounting for discrepancies between data sources would help overcome an inferred directionality remaining sensitive to sampling structure.

Our study design poses several limitations. First, mobility data were available only for 2020-2021, partially overlapping with the epidemiological sampling period. Second, viral spread was only indirectly linked to the full mobility network. While ethically justified, this metric does not explicitly account for other influencing factors such as fine-scale connectivity linked to public transport routes and use, movement associated with high-risk encounters and venues, or healthcare accessibility. Moreover, the lack of association between genetic distance and pairwise mobility reflects limitations in how mobility was quantified and could be improved using alternative metrics that better capture its effect on genetic structure. Finally, delays across the workflow from data generation to downstream analyses limit our capacity to inform public health decision-making in near real time. Thus, future efforts would benefit from continued collaboration between public health and academic sectors to enable faster responses.

Human mobility plays a central role in viral transmission in an increasingly connected world^42,43^. Our findings demonstrate that effective HIV control in large, densely populated urban areas must account for population-level movement alongside sustained investment in genomic surveillance. Although fine-scale dispersal patterns should be interpreted cautiously, the recurrent identification of highly connected boroughs and mobility hotspots may help prioritise geographically targeted prevention, surveillance, and healthcare interventions. However, translating these insights into public health impact will also require targeted prevention strategies that prioritise populations disproportionately affected by HIV, together with expanded surveillance in underrepresented areas of growing epidemiological relevance.

## METHODS

### Data access

HIV genotyping and sequencing procedures performed as part of the national drug resistance surveillance programme have been described previously^13,44^. Further details on HIV surveillance across Greater Mexico City are provided in **Supplementary Text 1.** The virus genetic data used here was generated by the Ministry of Health Mexico (CIENI-INER)^18^ and Clinica Especializada Condesa (CEC)^19^, with sequences corresponding to newly diagnosed cases reported between July 2019 and September 2022 mostly from Greater Mexico City, and the states of Chiapas and Quintana Roo (n = 9,338 partial and full polymerase gene sequences). The original dataset was filtered to retain only sequences from Greater Mexico City generated through CEC enrolment protocols, with length >2820 nucleotides, assigned to HIV-1 subtype B, and with residential postcode assigned. Thus, sequences generated through other CIENI sampling protocols within the study area and from neighbouring states (e.g., Hidalgo, Guerrero and Puebla) were excluded to maintain sampling consistency. This resulted in 7,078 sequences containing the following essential metadata: sampling location, residential postcode, biological sex (assigned at birth), sexual risk group classification incorporating gender identity, age, drug use (classified as category 1: all substances excluding methamphetamine; and category 2: methamphetamine use only), nationality and initial date of sample collection/diagnosis. All variable definitions for all metadata can be found in **Supplementary File 1 - Supplementary Table 2**. For sequence accession numbers, see **DATA AVAILABILITY** section.

### HIV transmission networks

The HIV transmission network across Greater Mexico City was reconstructed using the HIV-TRAnsmission Cluster Engine (HIV-TRACE)^22^. Global network statistics, including the number of nodes, edges, degree distribution, and cluster number and sizes were computed using the HIV-TRACE toolkit^22^ (www.hivtrace.org, www.github.com/veg/hivtrace). The pipeline first generates a codon-aware pairwise alignment relative to the reference sequence. The alignment is then inputted to compute pairwise genetic distances under the Tamura-Nei 93 (TN93) substitution model^45^, which performs well for large-scale datasets with low genetic diversity^22^. Pairwise distances were subsequently used to link sequence pairs under a specified genetic distance threshold. We further evaluated the impact of alignment length on genetic distance threshold selection and on cluster metrics (see **Supplementary Text 2** and **Supplementary Fig. 3**).

### Assortativity

We computed degree-weighted homophily (DWH) applying a heuristic scoring approach implemented in the Autotune tool^46^. DWH indices were computed based on four metadata attributes: sexual risk, methamphetamine use, nationality, and state of residence (CDMX or MEX) *(hivnetworkcsv -i path_outTN93.csv -f plain -A 0 > path_out_autotune_report.tsv).* For each attribute, a null distribution was generated by iteratively randomizing attribute labels across nodes and recomputing DWH scores. This permutation procedure defines a panmictic range corresponding to random mixing, against which observed DWH values were compared to identify non-random mixing^46^.

### Cluster growth

The Sr>2 metric was used to assess cluster growth. Sr>2 is defined as the number of newly diagnosed individuals joining a given transmission cluster within a defined time window (*e.g.,* the preceding 12 months), divided by the square root of cluster size at the end of the time window. This normalization accounts for differences in sizes, enabling a comparison of cluster growth rates^26^.

### Logistic regression

We fitted generalized linear models (GLMs) under a binomial distribution to test the following demographic variables as predictors of cluster membership and growth: sexual risk group, methamphetamine use, nationality and state of residence. Cluster membership was defined as a binary outcome (1 = in cluster, 0 = not in cluster). In parallel, we modelled cluster growth status (>2 cluster growing; <2, cluster not growing) as a binary outcome defined by the Sr>2 metric. Odds ratios (ORs) with corresponding 95% confidence intervals (CIs) were computed and reported with statistical significance assessed under a two-sided Wald test and a p-value of < 0.05. Missing data were handled using listwise deletion. Model fit was evaluated using the Akaike Information Criterion (AIC), whilst convergence was achieved using Fisher Scoring, requiring between 3 and 6 iterations across models. Analyses were applied to *i*) include all clusters within the network, and *ii*) restricted to the ten largest clusters identified.

### Epidemic growth

To assess epidemiological growth over time, we employed the BDSKY framework^47^ implemented in BEAST v2.7.5^48^ for estimating shared effective reproductive number (R_e_) and the rate of becoming uninfectious, with their corresponding 95% highest posterior density intervals (95% HPD). A hierarchical model was jointly applied to the 10 largest transmission clusters identified within the network, assuming shared population and evolutionary models (constant population size and a GTR+G substitution model). Analysis was done over three epochs: before 2017, between 2017 and March 2020, and after March 2020 until the end of the sampling period (2022). Priors used were as follows: for R_e_, we used a lognormal distribution with mean = 0 and standard deviation of 1 constrained between 0 and 7. For the rate of becoming uninfectious (δ), we applied a uniform prior constrained between 0.1 and 4 per year, corresponding to a plausible infectious period of 3 months to 10 years. Finally, the sampling proportion was fixed to 0 before the first sampled sequence in the analysis (July 2019) modelled under a beta distribution (α = 1, β = 1). The clock prior was fixed under a rate 1.15X10⁻^3^ s/s/y^49^. We ran two independent MCMC (Markov Chain Monte Carlo) chains for 200×10^6^ generations, removing the first 10% burn-in, and combining analysis-specific chains with *LogCombiner*^50^. R_e_ and δ estimates and associated 95% HPD are reported in **Supplementary File 1 - Supplementary Table 6**.

### Phylogenetic inference

The codon-aware pairwise alignment retrieved from HIV-TRACE was used for further phylogenetic inference. Phylogenetic trees were inferred using a maximum-likelihood (ML) framework using IQ-TREE multicore version 2.2.6 (*iqtree2 -nt 8 -m GTR+I+G -B 1000 -keep-ident*)^51^ (**Supplementary File 3**). Due to a lack of evolutionary rate estimates for the full 2.8 kb *pol* gene dataset, we selected a best-fitting rate identified under a credible TMRCA congruent with the 1.4 kb region. We tested seven fixed values ranging from 1.00 to 1.30X10⁻^3^ s/s/y, confirming that a rate of 1.15X10⁻^3^ s/s/y accurately recapitulates the TMRCA inferred when using the 1.4 kb data for HIV-1 subtype B circulating in North America^49^. The resulting ML tree was then time-calibrated using TreeTime^52^ (*treetime --aln X.fasta --tree X.nwk --dates X.csv --keep-root --stochastic-resolve –clock-rate 0.0015 --clock-filter 0*) informed by tip dates, constraining the tree to a rate specified above.

The CIENI-CEC sequences were further integrated within an extended dataset comprising near 40,000 partial HIV-1 B *pol* gene ‘background’ sequences collected between 2011 and 2022 from over 22 different countries in the Americas. According to availability from Los Alamos HIV database^53^, sequences were mostly sampled from Brazil, Mexico and United States with some representation from Argentina, Belize, Canada, Chile, Cuba, Ecuador, French Guiana, Guadeloupe, Guatemala, Honduras, Haiti, Martinique, Nicaragua, Panama, Peru, Puerto Rico, El Salvador, United States and Venezuela. Earlier-sampled sequences from previous CIENI-CEC protocols were also included, corresponding to other states in the country (excluding CDMX and MEX).

In order to minimize potential sampling biases in subsequent phylogeographic analyses^54^, an equal random number of ‘background’ sequences per year and region were subsampled. The filtered dataset comprised 37,384 pol sequences. The large-scale dataset was re-aligned using MAFFT v7.5^55^ (*mafft --auto --thread 8*) and inputted for phylogenetic inference using IQ-TREE multicore version 2.2.6^51^ (*iqtree2 -nt 8 -m GTR+I+G -B 1000 -keep-ident*). The resulting ML tree was time-calibrated informed by tip dates using TreeTime^52^, with outliers identified within the root-to-tip divergence plot iteratively removed, resulting in a final tree comprising 37,365 sequences including the CIENI-CEC data (**Supplementary File 4**).

### Discrete phylogeographic analysis

The ‘large-scale’ ML time tree was inputted as a fixed topology to perform phylogeographic analysis only considering two locations: Greater Mexico City and ‘Other’. For this purpose, we used the discrete trait analysis (DTA) implemented in BEAST v1.10.4^50,56^, and which employs an asymmetric substitution model to reconstruct the location states at ancestral nodes^57^. We ran a single Markov chain Monte Carlo (MCMC) until reaching chain convergence, inspecting effective sample sizes (ESS >200) for all relevant parameters using Tracer v1.7^58^. A maximum clade credibility (MCC) tree was then retrieved and annotated using TreeAnnotator^59^, whilst phylogeographic clades were annotated and visualized using FigTree v1.4.4. Tips were further annotated according to HIV-TRACE cluster, confirming correspondence between network-defined clusters and phylogenetic clades.

To assess the impact of potential sampling biases within our data relative to sampling locations (*i.e.,* 3:1 CDMX vs MEX), we generated three independent replica datasets comprising reduced versions of the large-scale dataset based on an epidemiologically-informed subsampling. Each replica dataset included all ‘background’ sequences and a fraction of the CIENI-CEC sequences (*n* = 1,896) randomly down-sampled according to the lowest sequences-to-incidence ratio per year per geographic location (**Supplementary File 1 - Supplementary Table 2**). Incidence was equivalent to number of new HIV cases per 100,000 inhabitants per year, as officially reported^21^. Independent DTAs were performed on each replica as described above.

### Continuous phylogeography and diffusion coefficients

Following the DTA, a continuous phylogeographic analysis^30^ was conducted *post hoc* at the clade level to investigate the dispersal dynamic of viral lineages introduced into the study area^56^. For this, we used the relaxed random walk diffusion model^60,61^ implemented in the software package BEAST 1.10^50^. The MCMC was run for 10^9^ iterations while sampling every 10^6^ iterations and discarding the first 10% sampled trees as burn-in. MCMC convergence and mixing properties were inspected with Tracer v1.7^58^ as described above. Geographic coordinates (latitude and longitude) were drawn from the Basic Geo-statistical Area (AGEB; Area Geoestadistica Basica) of origin of each sequence linked through assigned postcodes, retrieved from the official shapefile provided by the Mexican National Institute of Statistics and Geography (INEGI)^12,15^. AGEBs represent the highest resolution standardised geostatistical units in Mexico designed for the integration of demographic and socioeconomic information from national surveys, including population census^12^ (for validation analyses, see **Supplementary Text 4, Supplementary Fig. 12**).

Inferred lineage movements were projected onto a geo-referenced raster grid of Greater Mexico City (resolution = 0.05 arcmin). We used the R package “seraphim” v2.0^62,63^ to extract the spatio-temporal information embedded in spatially-annotated trees sampled from the posterior distribution, to then map the inferred dispersal history of viral lineages, and to estimate the weighted diffusion coefficient^31^ and isolation-by-distance (IBD) signal^29^ associated with each clade. The IBD signal was evaluated as the Pearson correlation between the patristic and log-transformed geographic distances between each pair of tip nodes within an introduced clade. We also investigate the association between the genetic (*i.e.* patristic) distance and our mobility-based metric (defined in the Methods section below). For global analyses combining all clades, Spearman’s rank correlation (ρ) was used to enable comparison between the IBD signal and our mobility-based metric.

### Mobility analyses

Human mobility data purchased from the company Veraset derived from anonymized mobile devices recorded daily nationwide between 01/01/2020 and 31/12/2021^15,32^. The source dataset comprised time-stamped, anonymised and aggregated mobile device observations associated with geographic coordinates, which were subsequently linked to AGEBs^12^. The full mobility network was reconstructed using directed, weighted origin-destination edges representing the mean normalized volume of observed trips between AGEBs across the study period. Second, AGEB-level flows were aggregated to define a municipality-level mobility network constrained to Greater Mexico City, with nodes corresponding to CDMX and MEX. In both representations, weighted and directed edges capture the intensity and directionality of population-level movement between locations. Mobility data were then indirectly linked to the CIENI-CEC sequence data by individually assigning postcodes to residential (“home”) AGEBs. Approximately 6% of sequences were excluded as they could not be assigned to AGEBs within the mobility network. The resulting cross-linked framework was used to derive the following metrics:

1. Mobility *(node-level mobility strength)* was defined as a measure of mobility strength at an AGEB level. To quantify this, we devised a total strength metric, defined as the sum of incoming and outgoing mobility strengths (the weights of influx + outflux edges). Values were normalized by the number of active devices per day to ensure comparability across days, aggregated per month, and summarized as the median across 24 months. No additional population-based normalization was applied, as AGEB-level population estimates can be missing or zero for some units (*i.e.,* rural AGEBs). This mobility strength metric was assigned to sequence pairs via their “home” AGEB and summarised for phylogeographic clades^29^.
2. Bidirectional travel volumes between CDMX and MEX were quantified using aggregated, directed edge weights representing the normalised volume of observed trips from CDMX to MEX and from MEX to CDMX across the full mobility network.
3. Geographic spread per transmission cluster was defined as the distribution of destination AGEBs reached from the set of “home” AGEBs associated with all sequences within a given cluster. AGEBs were ranked by the volume of mobility connections and summarized using quantile-based metrics.
4. Mobility hotspots were defined as the top 50 AGEBs observed over the full sampling period receiving the highest cumulative inbound mobility strength from all “home” AGEBs associated with the largest clusters within the transmission network, reflecting locations with the highest levels of trips or visits across the study period.

### Dataframes, Plots and Statistics

Logistic regression analyses were performed with the logit and associated statistical functions in R (v4.3.2) using the “stats”^64^ packages. For the IBD signal and mobility metrics, 95% highest posterior density (HPD) intervals were calculated using the “hdi” function in the “HDInterval” R package^65^. Ridgeline plots were generated with the “ridgeline” R package^66^. Tables were created in Microsoft Excel 2018. All visualizations and post-processing analyses were conducted using custom scripts in R/Python.

### Ethics Statement

The Ethics and Research Committees of the Instituto Nacional de Enfermedades Respiratorias Ismael Cosío Villegas (INER) gave ethical approval for this work (Project Codes E02-17 and E02-20). The Human Research Protection Program of the University of California San Diego (UCSD) gave additional ethical approval for this work (protocol number 190121). All participants provided written informed consent, and the study was conducted in accordance with the Declaration of Helsinki.

## DATA AVAILABILITY

The full Greater Mexico City-HIV transmission network is available in **Supplementary File 2** and can be interactively visualized using hivtrace-viz (https://github.com/veg/hivtrace-viz). The full HIV tree corresponding to the CIENI-CEC data from Greater Mexico City is available in **Supplementary File 3**. The full HIV tree corresponding to the ‘large-scale’ data is available in **Supplementary File 4**. Sequence accession numbers will be provided in **Supplementary File 5** upon publication of the manuscript and release of the embargo corresponding to the NCBI GenBank submission entry SUB16183816.

## ACKNOWLEDGMENTS

M.E.Z is funded by a UCL Rosetrees Excellence Fellowship UCL2024\2. MHR would like to thank the Fondo Conjunto de Cooperación México-Uruguay from the Agencia Uruguaya de Cooperación Internacional and the Agencia Mexicana de Cooperación Internacional para el Desarrollo (AMEXCID) for providing funding for the geospatial dataset. SD acknowledges support from the *Fonds National de la Recherche Scientifique* (F.R.S.-FNRS, Belgium), from the Research Foundation – Flanders (*Fonds voor Wetenschappelijk Onderzoek – Vlaanderen*, FWO, Belgium; grant n°G098321N), from the University of Brussels (ULB, Belgium) internal fund, and from the European Union Horizon 2020 project LEAPS (grant agreement n°101094685). T.I.V. is funded by the NIH NIDA R01DA057141. This project was supported in Mexico by funds from Secretaría de Ciencia, Humanidades, Tecnología e Innovación (SECIHTI) (grants PRONACE SALUD 303079 and CBF-2025-I-3513).

## Notes

### Competing Interest Statement

The authors have declared no competing interest.

### Author Declarations

The Ethics and Research Committees of the Instituto Nacional de Enfermedades Respiratorias Ismael Cosio Villegas (INER) gave ethical approval for this work (Project Codes 547 E02-17 and E02-20). The Human Research Protection Program of the University of California San Diego (UCSD) gave additional ethical approval for this work (protocol number 190121). All participants provided written informed consent, and the study was conducted in accordance with the Declaration of Helsinki.

