## Supplementary Information for "HIV Transmission Dynamics in Greater Mexico City are Shaped by Dense Spatial Mixing"

**CONTENTS**

**Supplementary Text 1. HIV surveillance in Mexico**
- **Supplementary Figure 1. Geographic context of the CIENI-CEC surveillance network**

**Supplementary Figure 2. Demographics of the HIV epidemic from the CIENI-CEC data**
**Supplementary Text 2. Sensitivity analysis for genetic distance threshold selection applied to the CIENI-CEC data**
- **Supplementary Figure 3. Genetic distance threshold for CIENI-CEC data**
- **Supplementary Figure 4. Cluster metrics across *pol* datasets**

**Supplementary Text 3. Extended Results**
**Supplementary Figure 5. Phylogenetic placement of the largest HIV-TRACE transmission clusters within the 'large-scale' ML tree**
**Supplementary Figure 6. Geographic mixing of clusters across sampling locations**
**Supplementary Figure 7. DTA-derived MCC tree**
**Supplementary Figure 8. Virus diffusion across clades including major transmission clusters**
**Supplementary Figure 9. Virus diffusion velocity compared to Isolation-By-Distance**
**Supplementary Figure 10. Geographic spread of the largest HIV transmission clusters inferred from human mobility**
**Supplementary Figure 11. Top 50 AGEBs representing mobility hotpots**
**Supplementary Text 4. Validation of AGEB- post code (CP) intersections**
- **Supplementary Figure 12. Comparison of spatial resolution between CPs and AGEBs**

**SUPPLEMENTARY FILES**

**Supplementary File 1 (Supplementary_Tables.xlsx)**
- **Supplementary Table 1. Dataset subsampling according to HIV incidence**
- **Supplementary Table 2. Dictionary of metadata variables**
- **Supplementary Table 3. Largest clusters within the network**
- **Supplementary Table 4. HIV-TRACE cluster correspondence with phylogenetic clades**
- **Supplementary Table 5. Assortativity relative to key demographic attributes**
- **Supplementary Table 6. Cluster Growth**
- **Supplementary Table 7. BDSKY results**

**Supplementary File 2 (HIV_Mexico_28KB_outNET_final.json)**
**Supplementary File 3 (HIV_CIENI-CEC.tree)**
**Supplementary File 4 (HIV_LARGE-SCALE.tree)**
**Supplementary File 5 (Accession_numbers.txt)**

### Supplementary Text 1. HIV surveillance in Mexico

Diagnostic screening of HIV has contributed to surveillance of the virus in Mexico since the early 2000's[1]. CEC comprises two specialized clinics in CDMX, located in the Cuauhtemoc and the Iztapalapa boroughs (**Supplementary Fig. 1**). CEC-Cuauhtemoc operates since 2000, whilst CEC-Iztapalapa opened in 2016 as a strategic expansion of HIV care[2]. CEC-Iztapalapa is located within the CDMX borough with the highest case counts, reflecting its strategic positioning at the boundary of CDMX to increasingly provide care to patients from MEX[2]. Following HIV diagnosis using the national diagnostic algorithm recommended by the Ministry of Health Mexico, comprising two presumptive tests and one confirmatory assay, full HIV 2.8 kb pol gene sequences are routinely generated, paired with anonymized, case-specific metadata collected through a consented questionnaire applied at the time of patient enrolment. Complementing official national epidemiological data for HIV[3,4], CIENI-CEC conducts baseline genotyping to monitor antiretroviral drug resistance[5]. Baseline genotyping entails routine monitoring of genetic markers associated with antiretroviral resistance to non-nucleoside reverse transcriptase inhibitors (NNRTIs)[5-7]. Standard sequencing protocols for HIV epidemiological surveillance worldwide typically target the first 1,497 nucleotides (1.4 kb) of the HIV polymerase gene, encompassing the protease (PR) and a portion of the reverse transcriptase gene (RT)[8], as mutations within these regions play a key role antiretroviral drugs resistance[9]. Nonetheless, a growing number of mutations associated with drug resistance have been detected within the integrase (IN) region, linked to an increased use of integrase inhibitors (INIs) as antiretroviral therapy (widely reviewed in Esposito et al[10]). In CIENI, sequencing of the complete viral pol gene comprising 2.8 kb was gradually introduced, and has since then has become the standard for CIENI-CEC protocols, as described in Davila-Conn et al. 2020[11].

### Supplementary Figure 1. Geographic context of the CIENI-CEC surveillance network

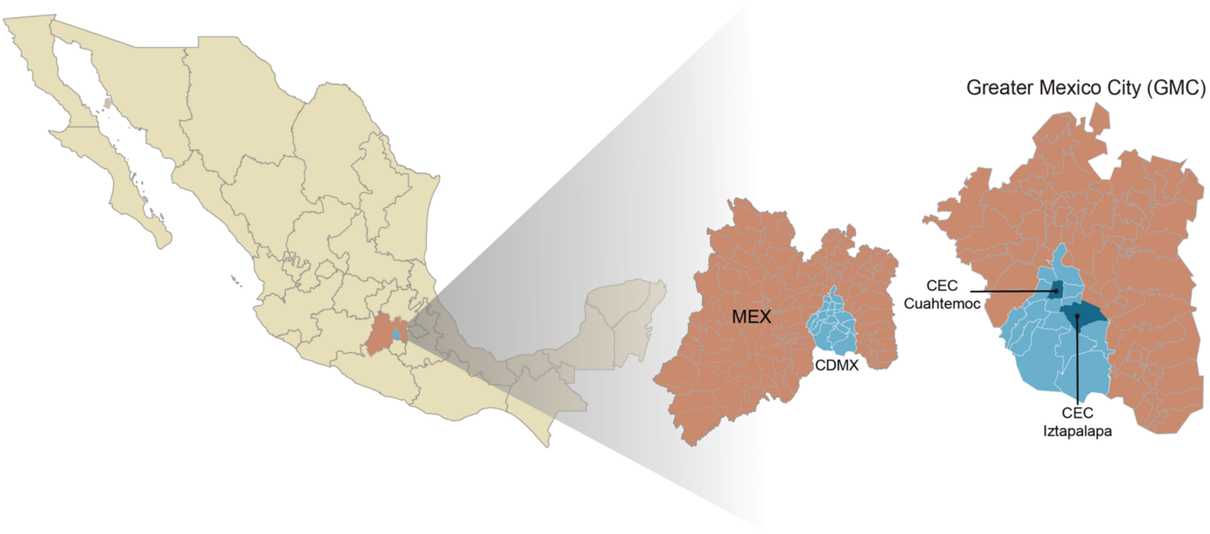


Geographic context of the CIENI-CEC HIV surveillance network within Greater Mexico City (GMC). The left panel shows Mexico (in yellow), showing MEX (highlighted red) and CDMX (in blue). Together, 16 CDMX boroughs and 59 adjacent municipalities from MEX comprise GMC. The right panel provides a zoomed view of GMC, showing CDMX and surrounding municipalities of MEX. Two CDMX boroughs host sister facilities comprising Clinica Especializada Condesa (CEC, indicated in dark blue): CEC-Cuauhtémoc (central CDMX) and CEC-Iztapalapa (eastern CDMX, adjacent to MEX). CEC centralises HIV diagnosis, care, and baseline sequencing for 50-70% of all new cases HIV reported in GMC. Map of GMC adapted from Wikimedia Commons. Public domain.

.

**Supplementary Figure 2. Demographics of the HIV epidemic from the CIENI-CEC data**

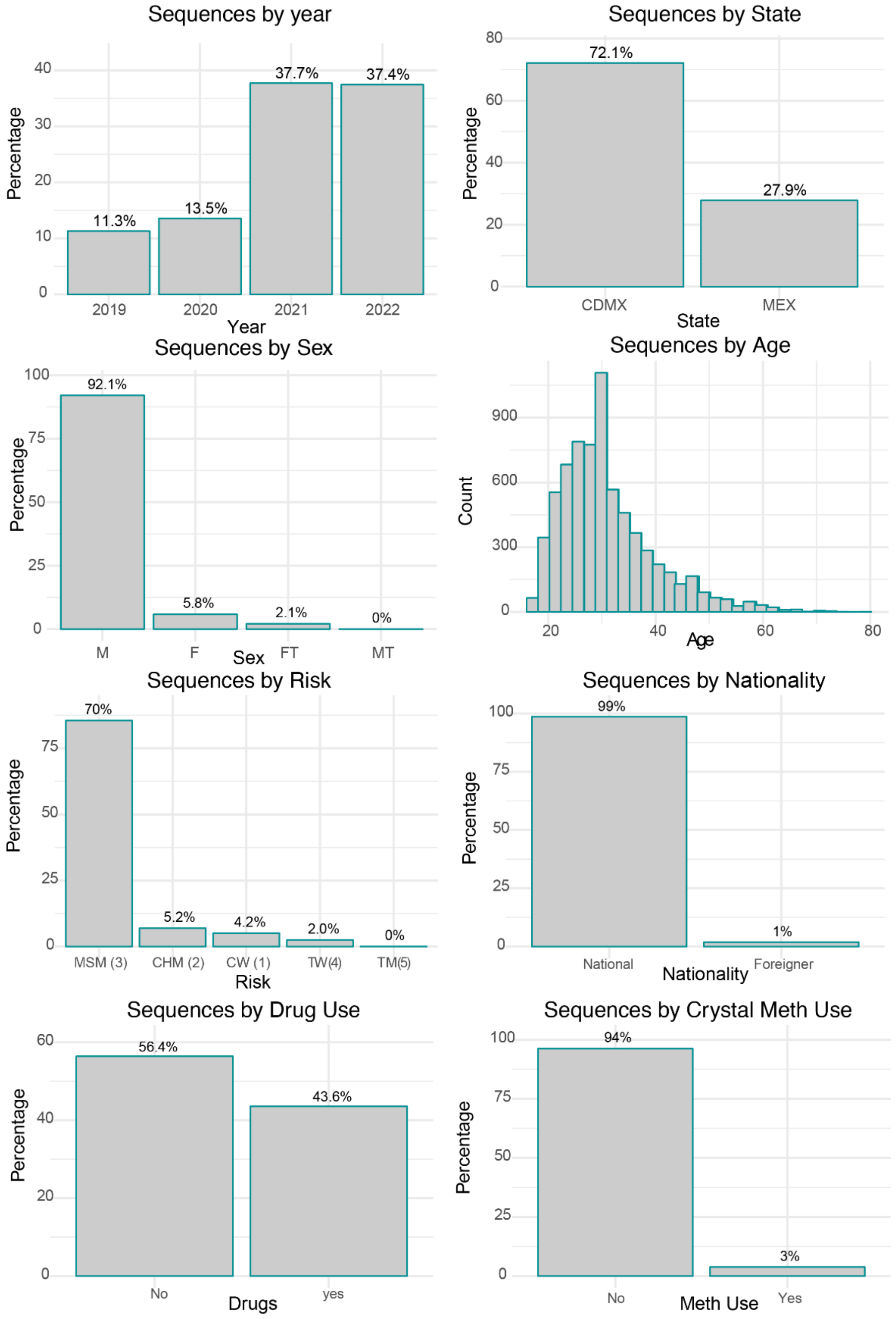


Histograms summarising the demographic structure of the CIENI-CEC data from GMC (7,078 *pol* gene sequences) by sampling year, geographic location, and demographic characteristics. Panels show the proportion of sequences by year of sampling (2019-2022), state of residency (CDMX/ MEX), sex assigned at birth (Female [F], Male [F]), age (displayed as a frequency distribution), sexual risk group classification incorporating gender identity (mem who have sex with men [MSM], cis heterosexual men [CHM], cis women [CW], trans women [TW], Non Available [NA],) nationality, drug use (all substances excluding methamphetamine use), and methamphetamine use only (Meth). Percentage values are shown above bars, where applicable.

**Supplementary Text 2. Sensitivity analysis for genetic distance threshold selection applied to the CIENI-CEC data**

The HIV-TRACE (TRAnsmission Cluster Engine) has been used to determine the optimal genetic distance threshold for the reconstruction of HIV transmission networks[12]. Based on global datasets comprising the partial 1.4 kb *pol* gene region of HIV-1 subtype B, a genetic distance threshold of 0.015 substitutions per site has been established for sequences with no more than 1.5% ambiguous positions relative to the HXB2 reference genome, a parameter defined as *ambiguity fraction*[12]. However, re-estimating an optimal genetic distance threshold applied to country-specific data has been recommended[13]. Selecting an appropriate genetic distance threshold tailored to the CIENI-CEC data was also important because the HIV-TRACE version tailored for internal use at CIENI-CEC (Seguro HIV-TRACE, https://seguro.hivtrace.org/) is configured with default parameters set for the partial 1.4 kb *pol* region[12]. Moreover, the added value of sequencing the full 2.8 kb *pol* gene for reconstructing transmission chains had not yet been determined prior to this work.

We performed a sensitivity analysis to identify a genetic distance threshold optimizing cluster formation applied to the CIENI-CEC data comprising the full 2.8 kb *pol* gene. To do so, we reconstructed multiple transmission networks across a range of genetic distance values (0.005-0.03 substitutions per site) using a fixed ambiguity fraction of 1.5% relative to the HXB2 HIV-1 subtype B reference genome[12] (GenBank accession: K03455). An optimal threshold close to 0.015 substitutions/site was estimated just before cluster formation decay is evident, indicated by a plateau in the curve (*i.e.,* when smaller clusters merge into larger, less informative clusters) (**Supplementary Fig. 3a**). We further evaluated the impact of alignment length on genetic distance threshold values. Under the expectation that a longer alignment (*i.e.,* higher genetic diversity) provides greater resolution for determining pairwise genetic distance, we expected that the distribution of sequence pairs would differ across datasets. This would result in incongruent values reflected by deviations from the x-y intercept in the regression plots. We performed individual analyses across partitioned alignments comprising different HIV pol gene regions: *i*) the full pol gene (2844 nucleotides, 2.8kb), *ii*) the replicase and partial integrase regions (1497 nucleotides, 1.4kb), *iii*) the *integrase* only (INT) region, and the *iv*) the non-linked (NL) pol sub-region. Sequence pairs across the 2.8kb and 1.4kb datasets show a strong correlation (slope= 0.99), with a x-y intersect value reflecting a congruent genetic distance threshold of 0.015 substitutions per site (**Supplementary Fig. 3b**).

Marginal differences in the number of clusters formed across all datasets were observed. Cluster numbers from the transmission network from the 2.8 kb data (n = 990) was comparable to those from 1.4 kb region data (n = 927) (**Supplementary Fig. 3a**). A notable difference was observed between the 1.4 kb and the INT region (n = 927 vs. n = 1,025, respectively). This observation is not surprising given the difference in length and functional properties of such gene regions, which may introduce greater variance when estimating pairwise distances. Cluster size distributions remained consistent across all data, with overlapping means in their respective cluster size distribution densities (range: 3.49-4.22) (**Supplementary Fig. 3b and c**). Overall, we find that alignment length has little impact on estimating genetic distance thresholds, as pairwise distances derived from the 1.4 kb and 2.8 kb pol alignments are highly correlated. This indicates that at an evolutionary scale relevant for transmission network reconstruction, full sequencing of the 2.8 kb *pol* gene increases precision but does not substantially impact the genetic distance threshold to that represents meaninfull epidemiological links.

**Supplementary Figure 3. Genetic distance threshold for CIENI-CEC data**

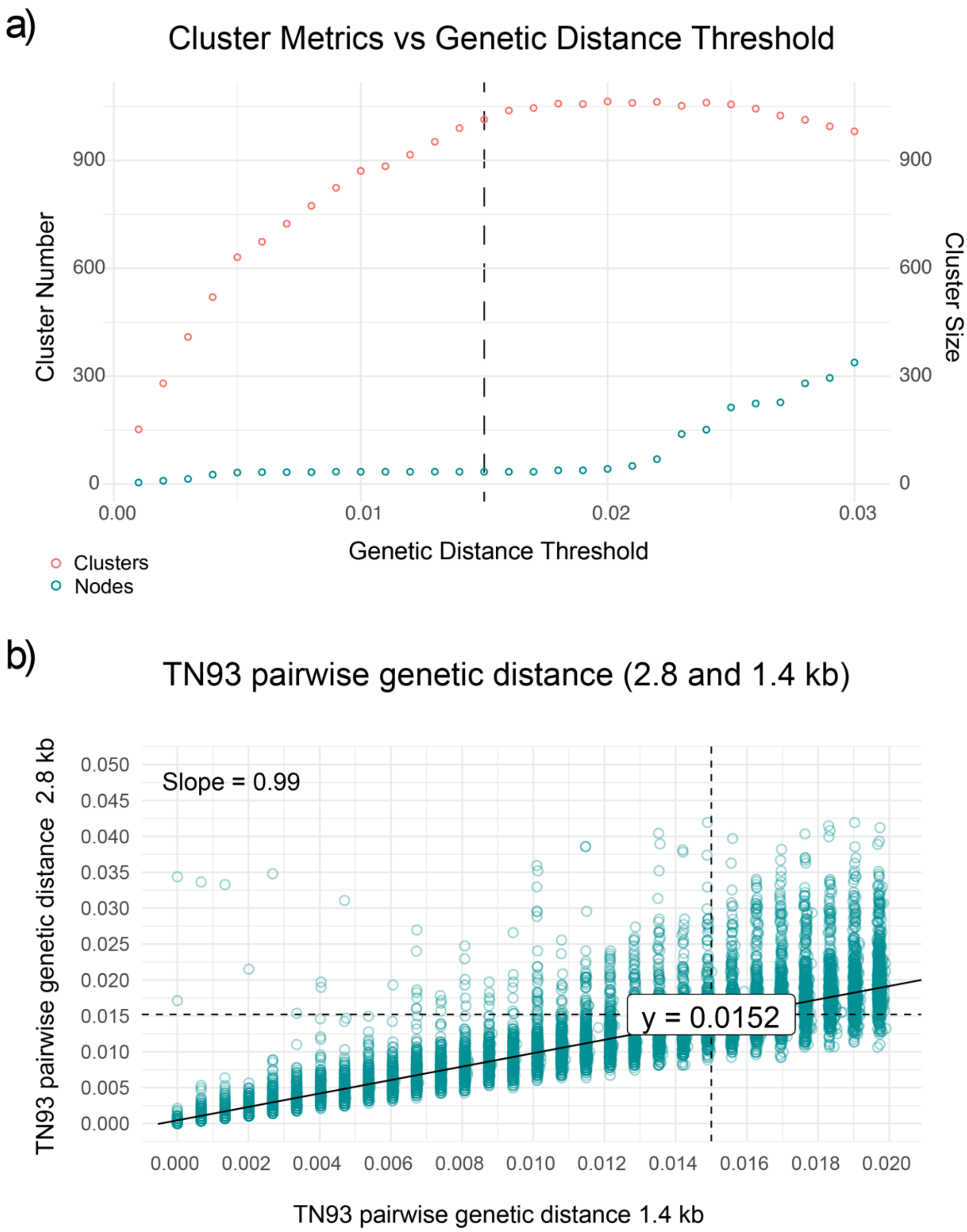


The relationship between genetic distance threshold and transmission network structure. a) the number of clusters (in red) and differences in size are reflected by number of nodes (in teal) for each transmission networks inferred across a range of genetic distance thresholds (0.005-0.03 subs/ per site). The vertical dashed line indicates the optimal threshold (~0.015 substitutions per site), corresponding to the point at which cluster formation begins to plateau, before smaller clusters merge into larger less informative clusters. b) comparison of pairwise TN93 genetic distances estimated from the full 2.8 kb and the partial 1.4 kb alignment. Each point in teal represents pairs of sequences with a given pairwise distance between them. The solid line shows the linear regression fit (slope = 0.99), indicating a strong correlation between distance estimates across alignments. Dashed horizontal and vertical lines mark the 0.015 substitutions per site threshold, demonstrating congruent threshold values (y-intercept ≈ 0.015).

**Supplementary Figure 4. Cluster metrics across *pol* datasets**

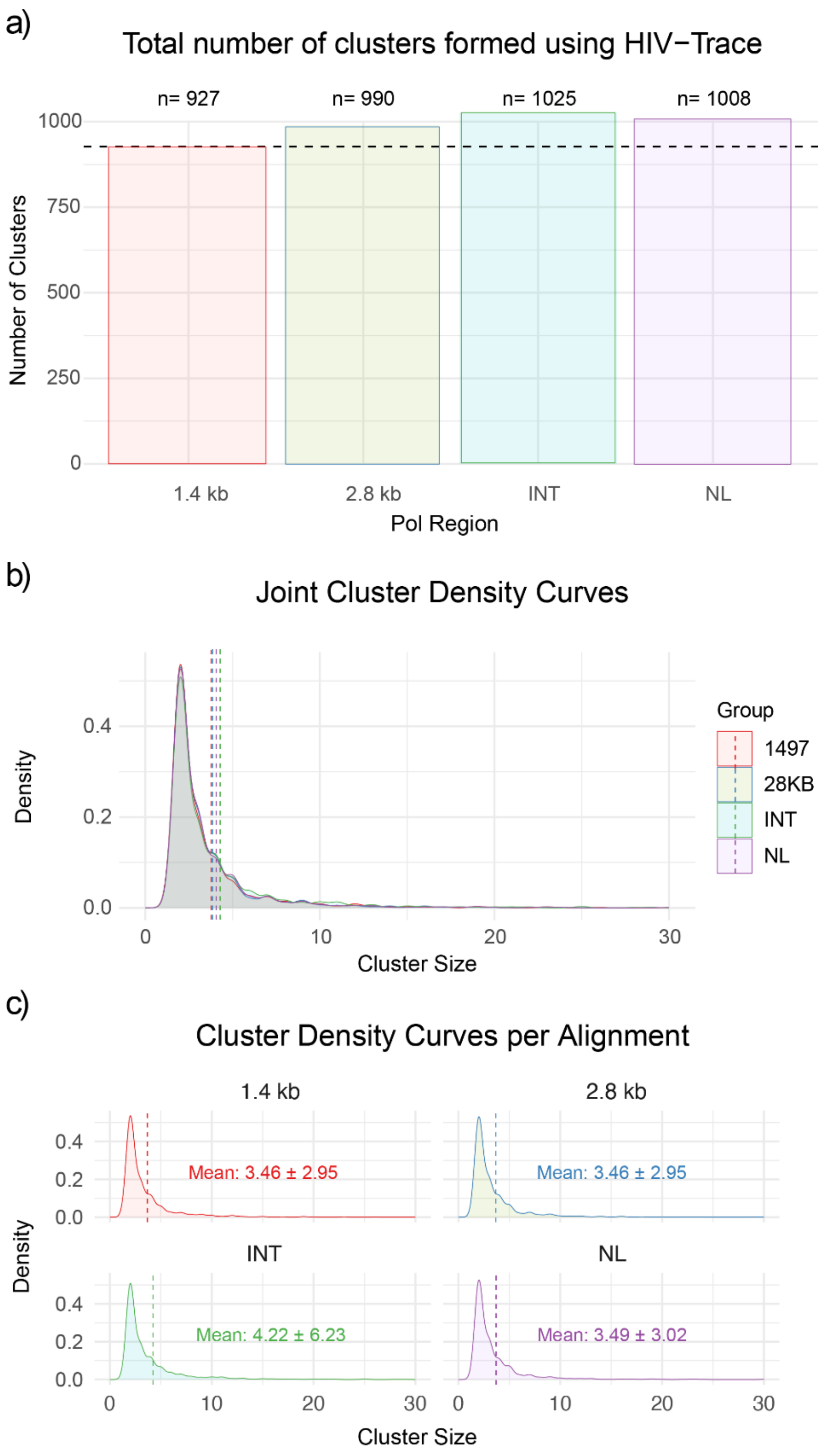


Total number of clusters across HIV-TRACE transmission networks. a) analysis was done across datasets representing different alignment lengths and *pol* gene regions: the partial 1.4 kb region, the full 2.8 kb gene, the integrase-only region (INT), and the non-linked region (NL). Numbers above bars indicate the total number of clusters formed the transmission network corresponding to each dataset. The dashed horizontal line highlights the cluster count inferred using the 1.4 kb pol region as a reference. b) Joint density distributions of cluster size distributions across datasets, showing overall similarity. Vertical dashed lines indicate the mean cluster size. c) cluster size density distributions shown separately for each dataset. The mean cluster size (± SD) are indicated within each panel, with overlapping distributions.

**Supplementary Figure 5. Phylogenetic placement of the largest HIV-TRACE transmission clusters within the 'large-scale' ML tree**

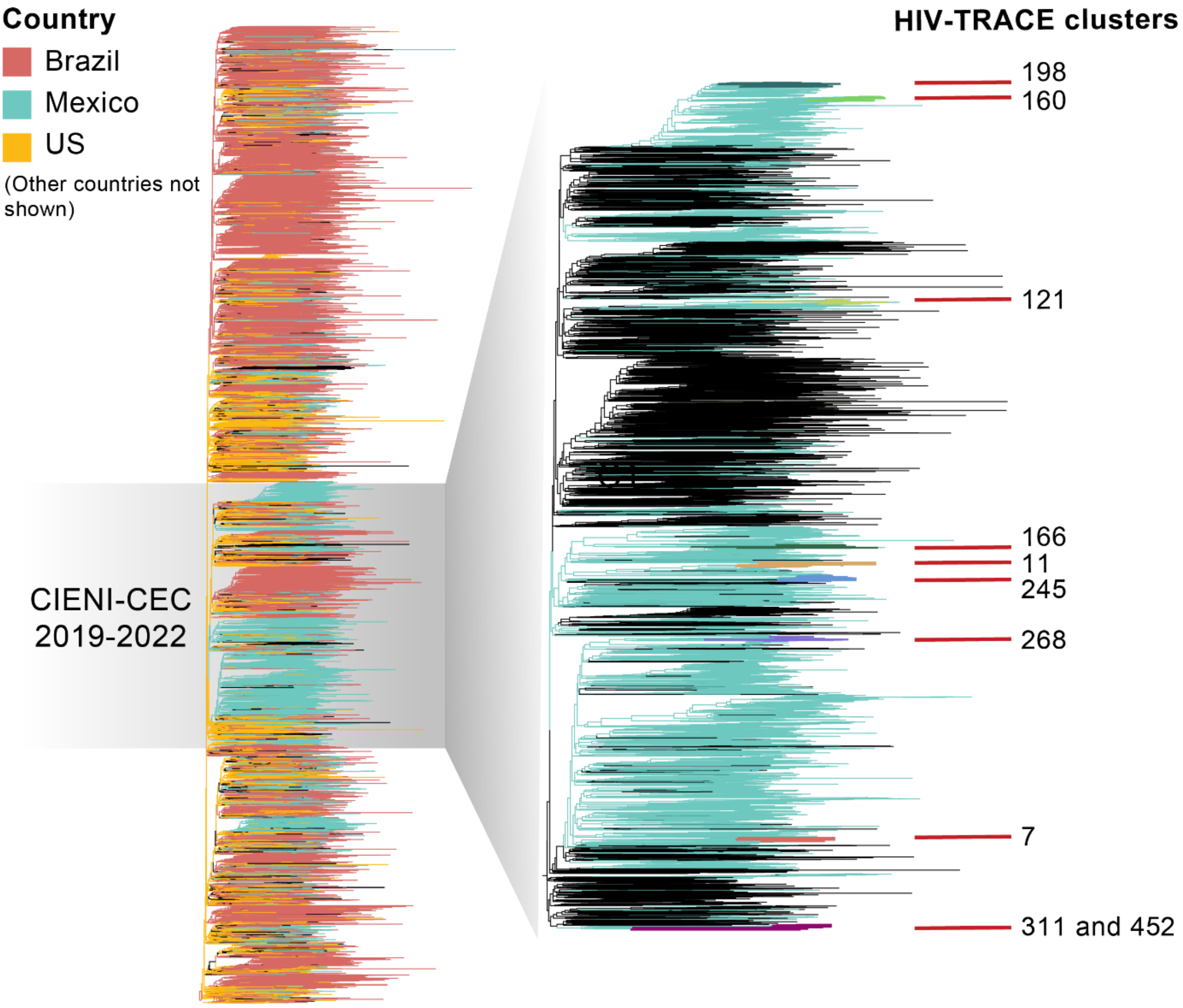


Maximum-likelihood (ML) phylogenetic tree inferred from the 'large-scale' dataset comprising 37,365 HIV-1 subtype B pol gene sequences sampled across 22 countries in the Americas between 2011 and 2022. The full tree is represented with branches coloured by sampling country (with Brazil, Mexico, and the United States shown; other countries with sparse sampling not shown for clarity). The clade corresponding to the CIENI-CEC dataset (7,078 sequences from GMC sampled between 2019 and 2022) is highlighted. On the right, a zoomed view of the monophyletic CIENI-CEC clade, showing the phylogenetic structure of locally circulating lineages, including all HIV-TRACE clusters. Branches corresponding to the ten largest HIV-TRACE transmission clusters are indicated with red labels and their corresponding ID. These correspond to monophyletic clusters within a broader phylogenetic context, confirming phylogenetic independence.

**Supplementary Figure 6. Geographic mixing of clusters across sampling locations**

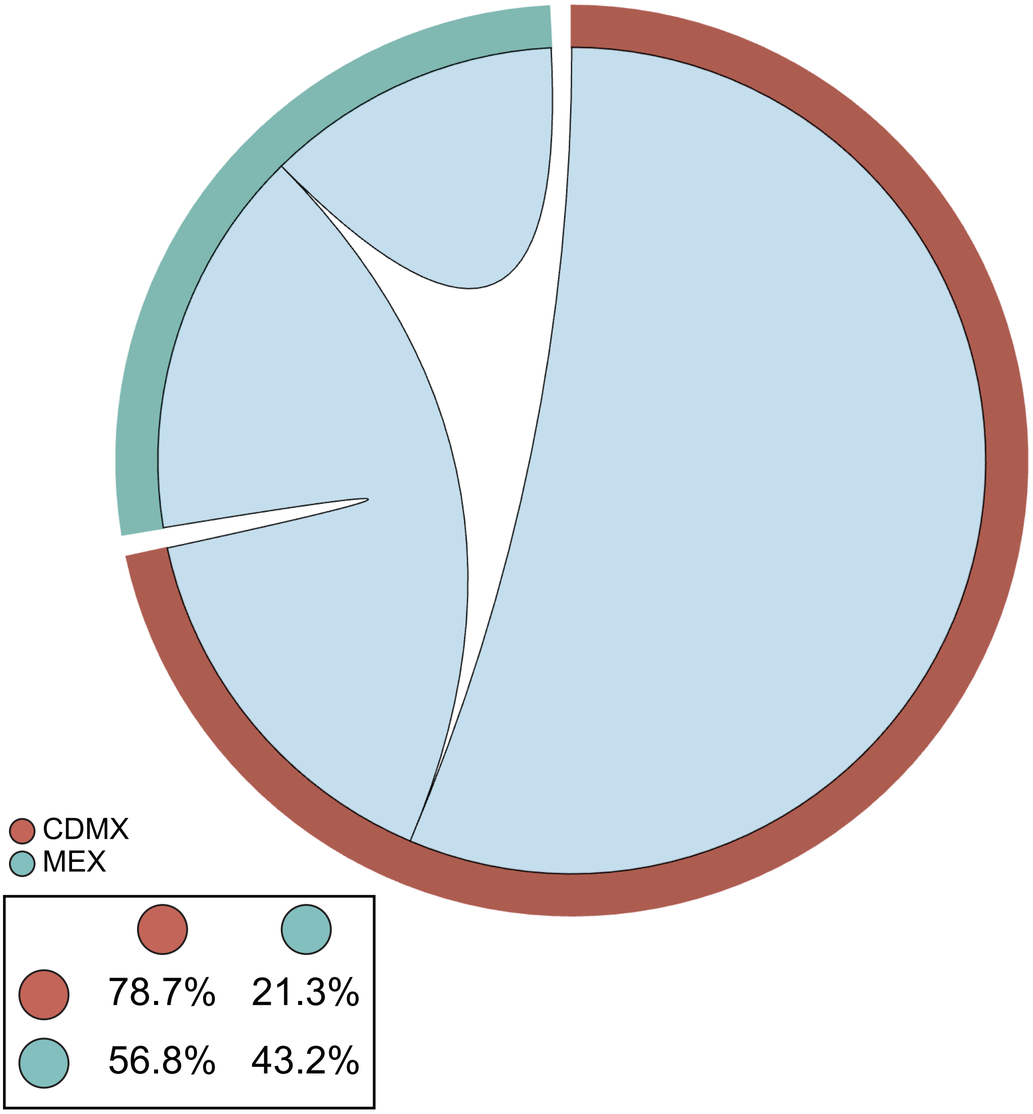


Chord diagram illustrating geographic mixing of clusters across sampling locations: CDMX (brown) and MEX (teal). Outer arcs represent the proportion of nodes originating from each location, while inner chords represent putative transmission links inferred from pairwise genetic distances. The inset shows the proportion of within-location and between-location connections stratified by location. For nodes from CDMX, most links occur within CDMX, with a smaller percentage of connections linking to MEX. In contrast, for nodes from MEX show a higher proportion of cross-location links, with the majority occurring within MEX and a smaller proportion linking to CDMX. This pattern indicate non-random geographic mixing and asymmetric connectivity across sampling locations.

**Supplementary Figure 7. DTA-derived MCC tree**

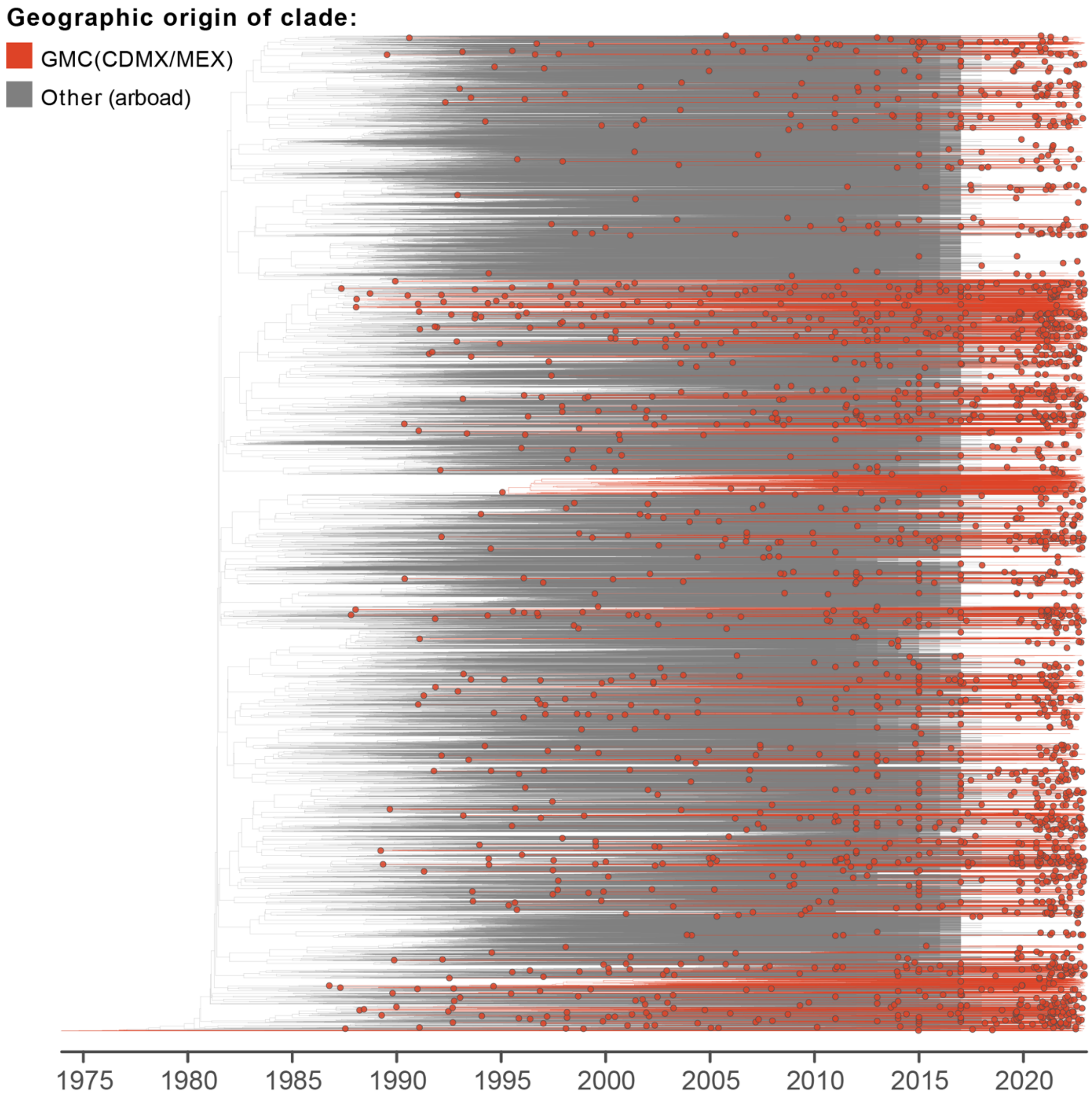


Maximum clade credibility (MCC) time-scaled tree inferred under a discrete phylogeographic analysis (DTA) using the 'large-scale' dataset comprising 37,365 HIV-1 subtype B pol gene sequences sampled across 22 countries in the Americas between 2011 and 2022, including all 7,078 CIENI-CEC sequences from GMC. Branches are coloured according to the inferred geographic origin of each clade: GMC (including CDMX/MEX, in red) and outside Mexico ("Other", in grey). Circles indicate posterior node state assignments for inferred geographic location of origin. We identified a total of 1,958 clades (≥3 nodes) corresponding to independent virus introduction events into Mexico from abroad.

**Supplementary Figure 8. Virus diffusion across clades including major transmission clusters**

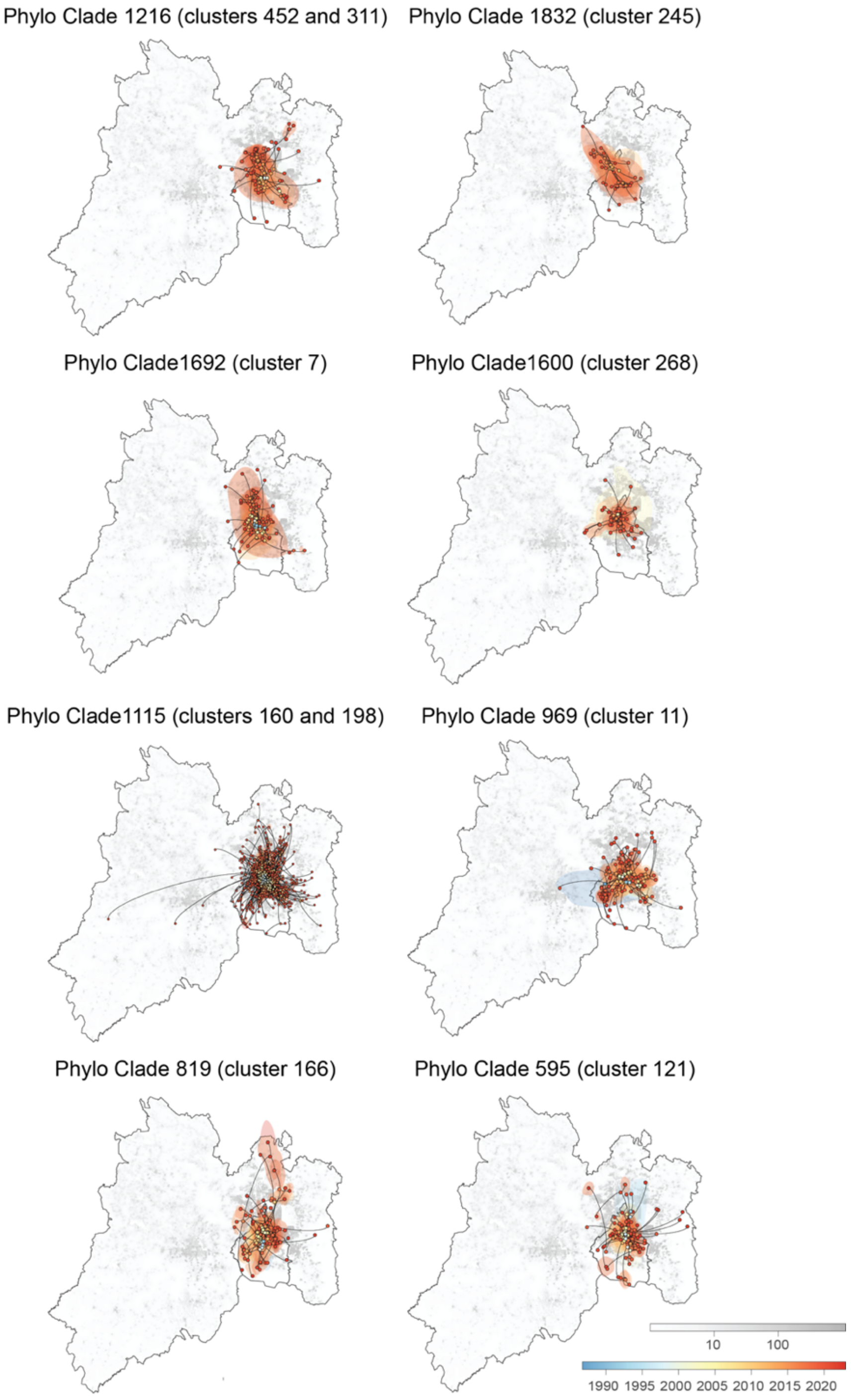


Map illustrating heterogeneous virus diffusion patterns across clades, ranging from relatively localised virus spread to broader spatial mixing across sampling locations. The reconstructed spatio-temporal virus diffusion patterns for selected clades containing the largest HIV-TRACE transmission clusters are shown. Map outlines delineate GMC, including CDMX and MEX. The grey-scale background represents log-transformed human population counts per raster cell. Points indicate inferred lineage locations through time, with colour ranges corresponding to sampling time according to the temporal scale shown. Trajectories were inferred under a relaxed random walk diffusion model on DTA-identified clades. Shaded polygons depict 80% highest posterior density (HPD) regions summarizing spatial uncertainty in lineage locations, estimated for one-year time slices.

### Supplementary Figure 9. Virus diffusion velocity compared to Isolation-By-Distance

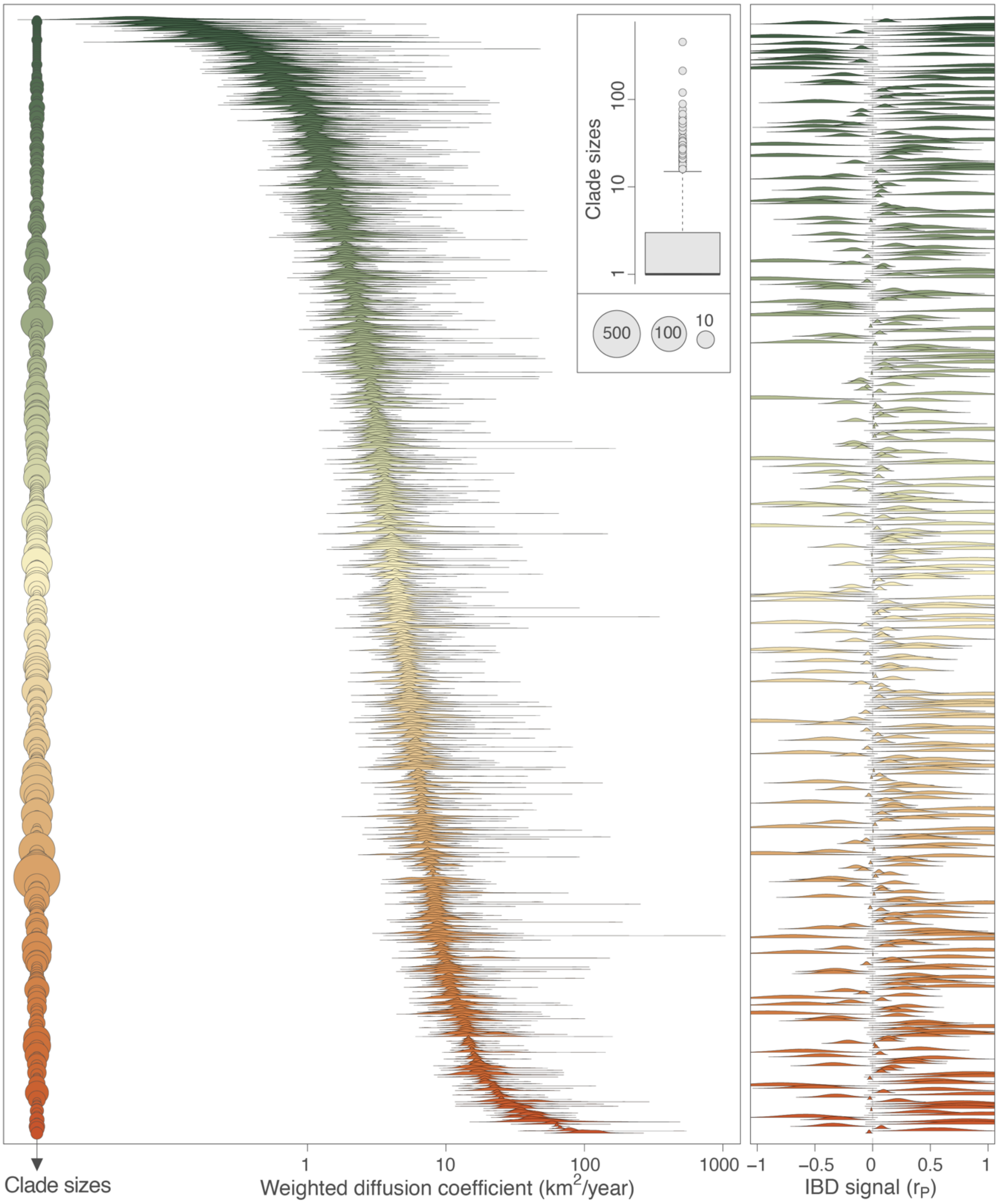


Clade-level lineage dispersal statistics inferred from the continuous phylogeographic reconstruction conducted for each phylogenetic clade (≥3 samples). Horizontal circular element represents single clades ordered by size (left), with point size proportional to the number of sequences within each. Colouring reflect relative clade size, from smaller (green) to larger (orange-red) clades. The weighted virus diffusion coefficient ($km^2$/year) represents the velocity of virus diffusion inferred under a relaxed random walk model. The right panel shows ridgeline density plots of the isolation-by-distance (IBD) signal, quantified as the Pearson correlation ($r_P$) between patristic and log-transformed geographic distances between pair of tip nodes within each clade. Each ridge corresponds to one clade and summarizes the distribution of its IBD signal estimate, centred on the dashed vertical line at $r_P$ = 0. Values close to zero indicate weak or absent distance-dependent spatial structure, whereas increasingly negative or positive values reflect stronger spatial structuring.

**Supplementary Text 3. Extended Results**

1. **Assortativity**

For most sexual risk groups, positive DWH values obtained (DWH = 0.024 risk group 1-CW, DWH = 0.016 risk group 3-MSM; DWH = 0.029 risk group 4-TW) indicate a weak assortative mixing by sexual risk group. In contrast, risk group 2-HCM showed negative DWH (-0.022) reflecting disassortative mixing. Estimates for risk group 5-TM were uninformative due to a small sample size. For methamphetamine use, DWH values were negative, again indicating disassortative mixing. Migratory status/nationality also showed no evidence of assortativity (**Supplementary File 1 - Supplementary Table 5**).

2. **Logistic regression**

Sexual risk groups 1-CW, 2-HCM and 4-TW had significantly lower odds for both cluster membership/growth compared to the reference group (3-MSM) (**Table 1**). Similarly, 'foreign' individuals had significantly lower odds of clustering relative to the reference category ('national'). Methamphetamine use is not a predictor for clustering/growth within the CIENI-CEC data. Residency location (CDMX vs MEX) is also not a predictor for clustering/growth. When analysis was applied to the ten largest clusters, overall patterns remained consistent. However, results for categories with small sample sizes should be interpreted cautiously, as small sample sizes may limit statistical power.

3. **Mobility between CDMX and MEX**

Short-term increases in mobility strength were observed during the initial COVID-19 lockdown period (March-May 2020); however, deviations were transient and did not result in a persistent directional shift or long-term reduction in metropolitan-scale movement[14,15]. From mid-2020 onward, bidirectional mobility was stable, with an isolated short-lived anomaly observed near the end of 2020. This anomaly likely reflects reporting or sampling artefacts rather than true behavioural change. A temporary skew favouring movement from MEX to CDMX is observed toward the end of the sampling period (October-December 2021). However, given the lack of mobility data after 2022, this pattern cannot be further investigated to be ruled at part of the expected variance. Several clusters exhibited broad spatial spread characterised by mobility connections between AGEBs spanning GMC, including northern municipalities of the MEX. Others displayed more spatially restricted patterns concentrated around central CDM

**Supplementary Figure 10. Geographic spread of the largest HIV transmission clusters inferred from human mobility**

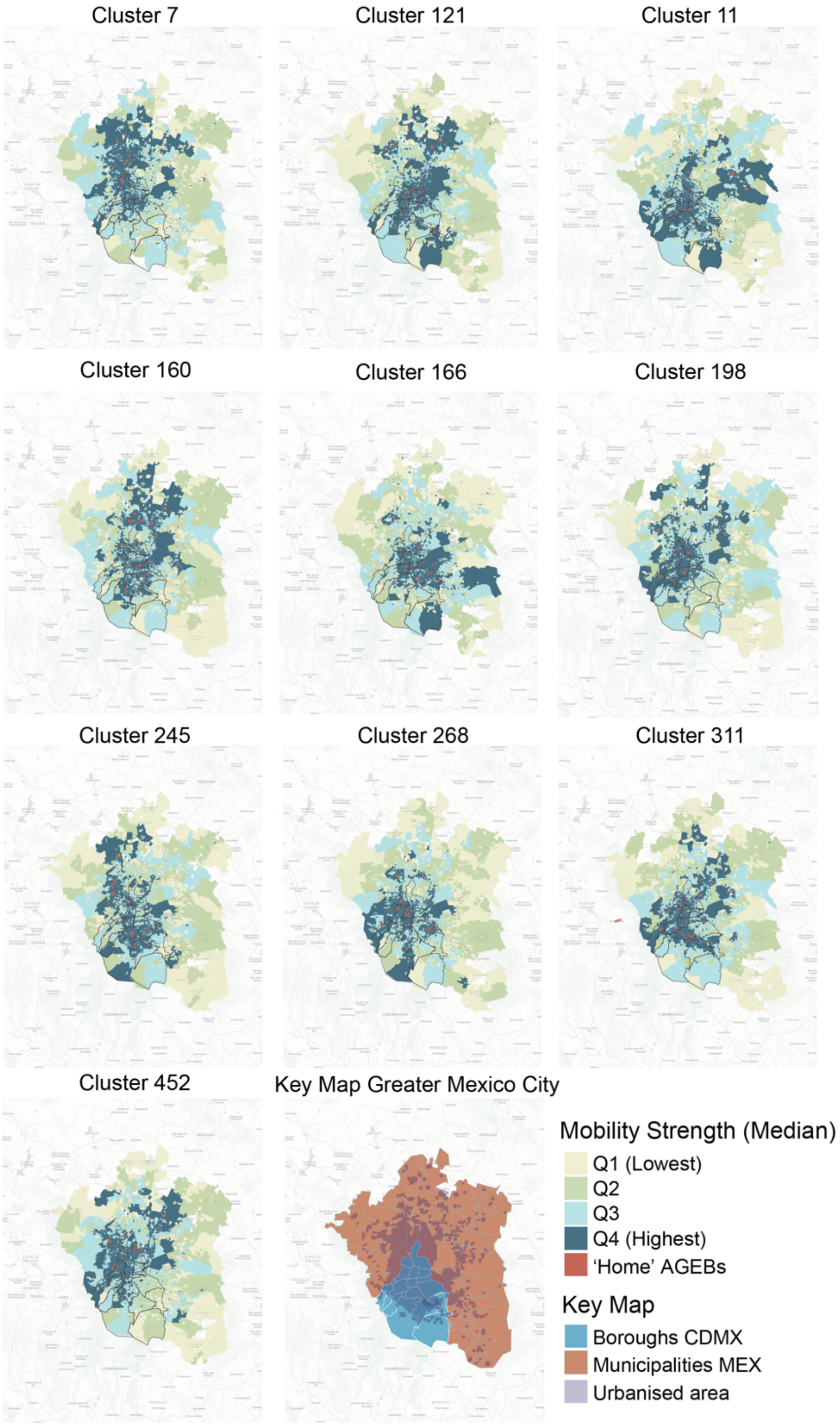


The set of residential ("home") AGEBs associated with all sequences within given clusters was linked to the full mobility network. Destination AGEBs reached from 'home' AGEBs were ranked according to the median volume of observed trips and summarized under quantiles (Q1-Q4), from lowest to highest mobility. Maps show the spatial distribution of 'home' and destination (visit) AGEBs for each cluster, with colour intensity reflecting increasing travel volumes. Borough boundaries for CDMX and for MEX municipalities are indicated for reference, together with the urbanised area of GMC.

**Supplementary Figure 11. Top 50 AGEBs representing mobility hotpots**

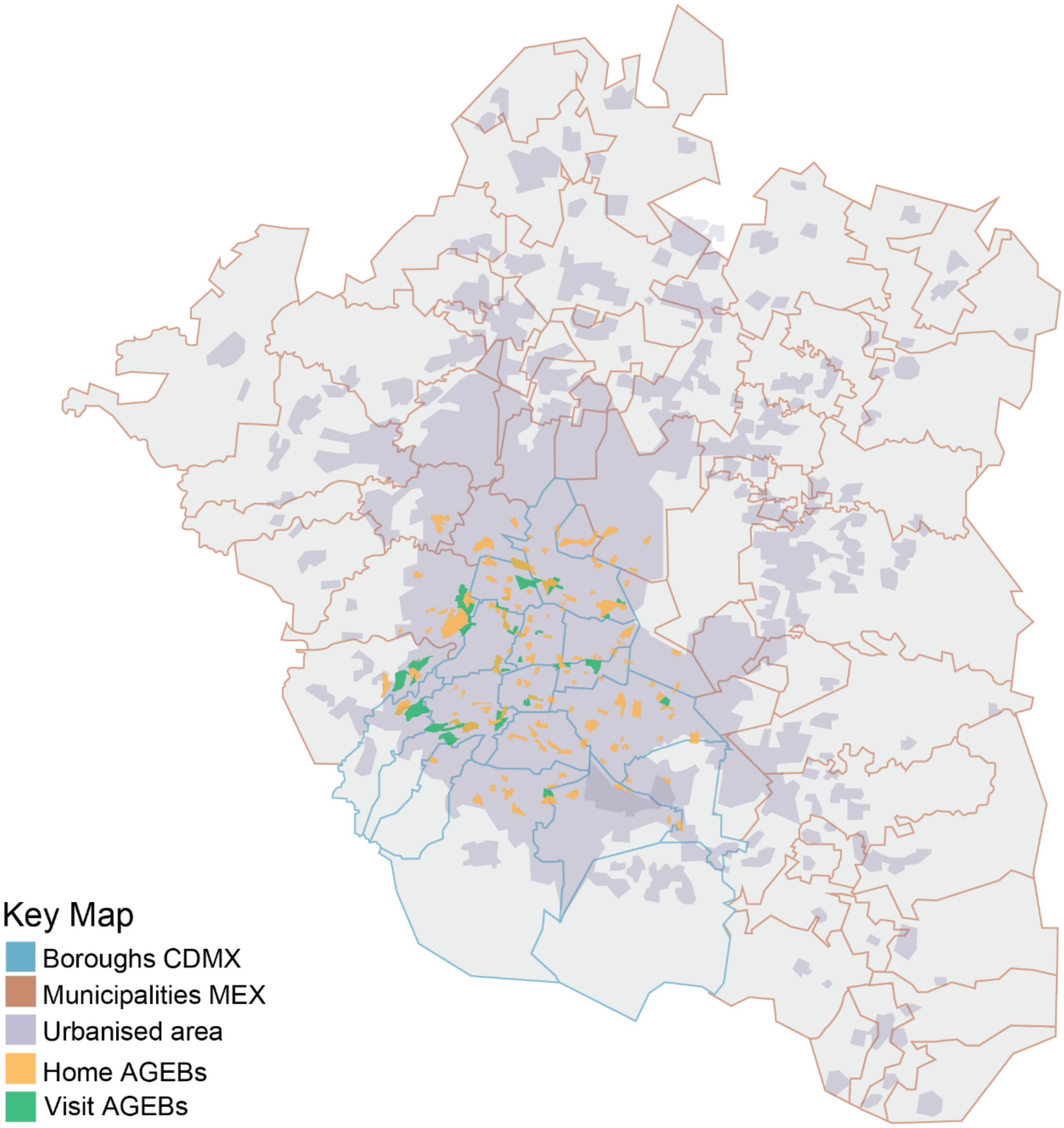


Map of GMC showing mobility hotspots. Blue boundaries delineate boroughs of CDMX, and pink boundaries indicate municipalities of MEX. Grey shading represents the urbanised area of GMC. Yellow polygons denote home (residential) AGEBs linked to cases within the largest clusters identified in the transmission network. Green polygons indicate mobility hotspots, defined as the top 50 AGEBs observed over the full sampling period that received the highest cumulative inbound mobility strength from all home AGEBs. The overlap between home and visit AGEBs highlights the concentration within the urban core of CDMX.

### Supplementary Text 4. Validation of AGEB- post code (CP) intersections

Analysis of 5,490 valid AGEB- post codes (CP) intersections revealed that based on geometric area, 70.5% of CPs were physically larger than the intersecting basic statistical areas (AGEB), while 29.5% of AGEBs exceeded their corresponding CPs in size. The same proportions were observed in the relative overlap metric, indicating that CPs not only cover larger ground areas but also tend to encompass a greater proportion of AGEBs. In urban areas, 77.5% of intersections showed CPs larger than AGEBs, reflecting the smaller, more densely subdivided AGEB units typical in urban areas. Conversely, in rural areas, only 12.3% of CPs were larger, as rural AGEBs generally encompass wider territories than their corresponding postal polygons. These observations highlight a scale mismatch between administrative (CP) and statistical (AGEB) boundaries, with AGEBs representing overall higher resolution. This is an important consideration for spatial epidemiological/mobility analyses in order to minimize aggregation bias and ensure accurate representation of population-level pattern

### Supplementary Figure 12. Comparison of spatial resolution between CPs and AGEBs

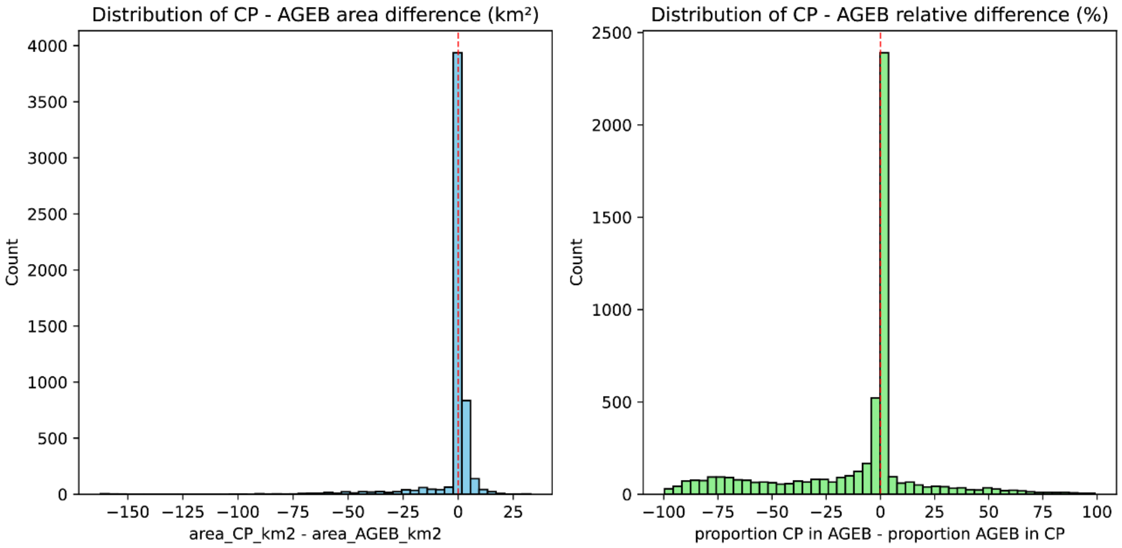


Histograms showing the distribution of (left) absolute area differences (CP - AGEB area, $km^2$) and (right) relative area differences, defined as the proportion of CP contained within AGEB minus the proportion of AGEB contained within CP. Positive values indicate cases in which CP polygons are larger or cover a greater fraction of the intersecting AGEB than vice versa. Vertical dashed lines denote zero difference. Most intersections are skewed toward positive values in both metrics, indicating that CPs generally span larger spatial extents and encompass a greater proportion of AGEBs, particularly in urban settings. The distributions highlight a systematic scale mismatch between administrative post code units and census-based statistical units, with AGEBs providing higher spatial resolution for mobility and epidemiological analyses.
